# An investigation of spatial-temporal patterns and predictions of the COVID-19 pandemic in Colombia, 2020-2021

**DOI:** 10.1101/2021.07.28.21261212

**Authors:** Amna Tariq, Tsira Chakhaia, Sushma Dahal, Alexander Ewing, Xinyi Hua, Sylvia K. Ofori, Olaseni Prince, Argita Salindri, Ayotomiwa Ezekiel Adeniyi, Juan M. Banda, Pavel Skums, Ruiyan Luo, Leidy Y. Lara-Díaz, Raimund Bürger, Isaac Chun-Hai Fung, Eunha Shim, Alexander Kirpich, Anuj Srivastava, Gerardo Chowell

## Abstract

Colombia announced the first case of severe acute respiratory syndrome coronavirus 2 on March 6, 2020. Since then, the country has reported a total of 4,240,982 cases and 106,544 deaths as of June 30, 2021. This motivates an investigation of the SARS-CoV-2 transmission dynamics at the national and regional level using case incidence data. Mathematical models are employed to estimate the transmission potential and perform short-term forecasts of the COVID-19 epidemic trajectory in Colombia. Furthermore, geographic heterogeneity of COVID-19 in Colombia is examined along with the analysis of mobility and social media trends, showing that the increase in mobility in July 2020 and January 2021 were correlated with surges in case incidence. The estimation of national and regional reproduction numbers shows sustained disease transmission during the early phase of the pandemic, exhibiting sub-exponential growth dynamics. Moreover, most recent estimates of reproduction number are >1.0 at the national and regional levels as of May 30, 2021. Further, the 30-day ahead short-term forecasts obtained from Richards model present a sustained decline in case counts in contrast to the sub-epidemic and GLM model. Nevertheless, our spatial analysis in Colombia shows distinct variations in incidence rate patterns across different departments that can be grouped into four distinct clusters. Lastly, the correlation of social media trends and adherence to social distancing measures is observed by the fact that a spike in the number of tweets indicating the stay-at-home orders was observed in November 2020 when the case incidence had already plateaued.

**Author summary:** As the COVID-19 pandemic continues to spread across Colombia, studies highlighting the intensity of the pandemic become imperative for appropriate resource allocation and informing public health policies. In this study we utilize mathematical models to infer the transmission dynamics of SARS-CoV-2 at the regional and national level as well as short-term forecast the COVID-19 epidemic trajectory. Moreover, we examine the geographic heterogeneity of the COVID-19 case incidence in Colombia along with the analysis of mobility and social media trends in relation to the observed COVID-19 case incidence in the country. The estimates of reproduction numbers at the national and regional level show sustained disease transmission as of May 30, 2021. Moreover, the 30-day ahead short-term forecasts for the most recent time-period (June 1-June 30, 2021) generated from the mathematical models needs to be interpreted with caution as the Richards model point towards a sustained decline in case incidence contrary to the GLM and sub-epidemic wave model. Nevertheless, the spatial analysis in Colombia shows distinct variations in incidence rate patterns across different departments that can be grouped into four distinct clusters. Lastly, the social media and mobility trends explain the occurrence of case resurgences over the time.

## Introduction

Ever since the detection of the first cluster of severe acute respiratory syndrome coronavirus 2 (SARS-CoV-2) in December 2019, the coronavirus disease 2019 (COVID-19) pandemic continues to threaten the world [1, 2]. This pandemic has been the defining global health event of the twenty-first century, resulting in devastating global socioeconomic and political consequences [3]. The virus’s novelty and complex transmission dynamics have resulted in non-pharmaceutical public health measures including social distancing mandates and intermittent lockdowns as the primary weapons to fight COVID-19 [4]. Moreover, as new variants of SARS-CoV-2 continue to emerge amidst vaccination campaigns globally, it remains unclear how the COVID-19 pandemic will unfold [5, 6]. Despite the uncertainty in the development of the pandemic, SARS-CoV-2 continues to exert substantial morbidity and mortality burden surpassing 180 million confirmed cases and more than 4 million associated deaths worldwide as of June 30, 2021 [7]. The COVID-19 mortality and morbidity surges continue to overwhelm the health care systems of many nations including Indonesia, Colombia, Mexico and Argentina [8–10].

It is imperative to understand the dynamics of disease transmission and the potential role of mitigation strategies at different spatial scales (e.g., municipality, regional or departmental level) in order to contain the virus [11]. Assessing the geographical patterns of the cases and identifying the COVID-19 impact at the regional level can provide valuable insights about the exposure to the virus and the distribution of public health resources required to mitigate its spread [12, 13]. Pandemic preparedness including the implementation and lifting of dynamic lockdowns [14], social distancing, mask mandates, population testing, and provision of vaccinations has varied across countries in the same region and in different cities within the same country [15]. As with Latin America, the evolution of the pandemic and the quality of public health response have varied across nations. While Chile aimed at mitigating the virus (slow down the disease and reduce peak in health care demand), Brazil, Ecuador and Colombia’s objective was to contain the spread of the virus (minimize the risk of transmission from infected to non-infected individuals to stop the outbreak) [16, 17]. Despite aiming to contain the viral transmission in the country, Colombia became the second country in Latin America and eighth globally to reach one million cases by the end of October 2020 [18]. Colombia has also seen the fastest increase in total COVID-19 associated deaths compared to other Latin American countries by the end of the year 2020 [16].

In Latin America, where the first confirmed COVID-19 case was reported in Brazil on February 26, 2020 [19], the intensity and diversity of social distancing efforts have varied widely. As the virus spread across Latin America, Colombia declared its first imported confirmed case on March 6, 2020 [20]. As the virus disseminated throughout the country, initial prevention recommendations (provided between March 9-11, 2020) from the local government authorities conflicted with those at the national level [21]. Consequently, the public was given vague preventive measures from health authorities [21]. Therefore, the case incidence continued to rise. This development led the authorities to close schools and universities on March 11, 2020, as a part of the prompt escalation of measures to prevent a full-blown outbreak [22]. This action was followed by the closure of the country borders on March 16, 2020, and Colombia declared a national emergency the next day [23, 24].

As cases kept increasing rapidly, domestic and international flights were suspended on March 23, 2020 [21]. Subsequently, a nationwide mandatory quarantine was implemented on March 24, 2020, to limit virus transmission within the country. Meanwhile a phased reopening of the economy was initiated as early as April 27, 2020, under strict protocols to support the country’s dwindling economy [25].

The mandatory quarantine was extended five times during a period of five months and lasted until August 31, 2020 [26, 27]. At this point, the country transitioned into a period of “selective isolation with responsible individual distancing” as the daily incidence in the country’s main cities including Bogotá, Medellín, Cali, Bucaramanga and Pasto levelled off and eventually leaned towards a downward trend [27]. Moreover, Barranquilla, Cartagena, Leticia and Quibdó had overcome the worst part of the case incidence trajectory by August 25, 2020 [27]. The chosen selective isolation strategy prioritized tracing of suspected cases, those with infection, and their contacts, while reactivating the social and economic life of the country. The government imposed and lifted the dynamic lockdowns in multiple cities across the country as the cases continued to increase in the subsequent months. By the end of year 2020, most of the COVID-19 cases were concentrated in capital city Bogotá (314,745), followed by Antioquia (163,952), and Valle del Cauca (81,101) [28]. As the pandemic entered its second year in Colombia, the P.1 (Gamma) variant of SARS-CoV-2 initially circulating in Brazil was detected on January 30, 2021. Soon after, the government announced a mass vaccination strategy that began on February 20, 2021 [29]. Around the same time, on February 25, 2021, the president extended the duration of national health emergency to last until May 31, 2021 [30]. Despite the administration of vaccines in the country, the cases kept rising in some cities like Barranquilla. This resulted in the implementation of curfews in the cities and municipalities based on the intensive care unit (ICU) occupancy rates in the hospitals beginning March 25, 2021 [31]. The implementation and lifting of curfew followed by the opening of businesses has continued in cities depending on the ICU occupancy rates of the hospitals till date. While the impact of COVID-19 pandemic was not uniform across the entire country, Bogotá, Cali and Cartagena have been the hardest hit areas in Colombia [32]. Despite being one of the first countries in South America to offer the diagnostic tests for COVID-19 [33], the testing rate for Colombia remains low as of June 30, 2021, with 2.15 tests per thousand people per day and a ∼31.4% positivity rate [34].

The Colombian government’s response to contain the COVID-19 pandemic has been declared the third worst out of 116 countries evaluated by the Lowy Institute in Australia. While the rank of Colombia’s response towards the COVID-19 pandemic is only ahead of Mexico and Peru [35], the country stands with a total of 4,240,982 cases and 106,544 deaths as of June 30, 2021 [36]. Three distinguishable events have particularly added to the complexity of the COVID-19 pandemic in Colombia. These include the outbreaks in prisons and nursing homes [37, 38] and the 2021 Colombian protests that started on April 28, 2021, against the tax reforms [30]. The COVID-19 pandemic severely impacted Colombian prisons in Cali, Villavicencio, and Bogotá due to over-crowding, inadequate medical supplies and unhygienic conditions of facilities, which led to many infected inmates [38]. Moreover, the COVID-19 outbreak in the nursing home in Manizales in August 2020 resulted in 74 elderly testing positive for SARS-CoV-2 [37]. Both these events affected the vulnerable sectors of the society across different departments in the country.

As the epidemic trajectory of the COVID-19 pandemic continues to unfold in Colombia it is vital to assess the future of the COVID-19 pandemic in the country amidst the interventions to control the spread of virus in the country. In order to forecast the epidemic trajectory of SARS-CoV-2, we utilize mathematical models that have been validated for previous infectious disease outbreaks such as Ebola and Zika [39, 40]. We also explore the spatial epidemic curves that allow us to analyze geographic heterogeneity that is lost while evaluating the epidemic curves at coarser scales. Specifically, we investigate the transmission dynamics of SARS-CoV-2 at the national and regional levels utilizing the case incidence data by the dates of symptom onset. We estimate the effective reproduction numbers of SARS-CoV-2 at the regional and national level and examine the social media and mobility trends concerning the implementation of lockdowns.

## Methods

### Data

Five sources of data are analyzed in this study. A brief description of the data sets is provided as follows:

(i) Case incidence data

Case incidence data by the dates of symptom onset is used to generate short-term forecasts of the COVID-19 pandemic, assess the spatial heterogeneity in Colombia and estimate the national and regional reproduction numbers. Specifically, we utilize publicly available time series of laboratory-confirmed cases by dates of symptom onset for the national and regional COVID-19 case incidence. These data were retrieved from the Colombian Ministry of Health as of June 30, 2021 [41]. Colombian departments and their four main districts; Barranquilla, Bogotá, Cartagena, and Santa Marta can be grouped into five regions as follows: the Amazon, the Andean, the Orinoquía, the Caribbean and the Pacific [42]. To evaluate the spatial heterogeneity and estimate the regional reproduction numbers, for simplicity we assume a regional level distribution as follows:

<u>Amazon region</u>: Amazonas, Caquetá, Guainía, Guaviare, Putumayo and Vaupes.

<u>Andean region</u>: Antioquia, Boyacá, Bogota, Caldas, Cauca, Cundinamarca, Huila, Nariño, Norte De Santander, Quindío, Risaralda, Santander, Tolima.

<u>Orinoquía region</u>: Arauca, Casanare, Meta, Vichada

<u>Caribbean region</u>: Atlántico, Bolivar, Cesar, Cordoba, La Guajira, Magdalena, San Andres, Sucre, Barranquilla, Cartagena, and Santa Marta

<u>Pacific region</u>: Choco, Nariño, Valle Del Cauca

Based on the geographical distribution of the departments, Nariño, Valle Del Cauca, Guainia and Cauca appear in two regions each. However, we include them in one region as stated above, to avoid the duplication of data for our analysis.

(ii) Genomic data

To estimate the reproduction number from the genomic data, 136 SARS-CoV-2 genome samples for Colombia (from the 1855 samples available as of June 30, 2021) were obtained from the “global initiative on sharing avian influenza data” (GISAID) repository [43] between February 27 and April 5, 2020 to estimate the early reproduction number.

(iii) Mobility trends data

Two sources of mobility data are analyzed at the national level from March 6, 2020 to June 30, 2021.

Apple’s mobility data is generated by counting the number of requests made to Apple Maps for directions in select countries/regions, sub-regions, and cities. This data is published publicly by Apple’s mobility trends reports and was retrieved as of June 30, 2021 for Colombia [44]. This aggregated and anonymized data are updated daily and includes the relative volume of directions requests per country compared to the baseline volume on January 13, 2020. Apple has released the data for the three modes of human mobility: driving, walking and public transit. The mobility measures are normalized in the range 0-100 for each country at the beginning of the series; thus trends are relative to this baseline.

Google’s mobility data [45] is retrieved from the users who have turned on the location history in their Google account. Google’s mobility data shows the changes in mobility in each geographic and administrative region for visits to different places, such as grocery stores, parks, and recreation spots. The baseline day represents the normal value for the day of the week. The baseline day is the median value from the 5-week period from January 3, 2020 to February 6, 2020.

(iv) Twitter data

For Twitter data analysis, we retrieved data from the publicly available Twitter data set of COVID-19 chatter from March 12, 2020 to June 30, 2021 [46]. A detailed description about Twitter data acquisition and processing is provided in a prior study [46].

### Modeling framework for forecast generation

We utilize the dynamic phenomenological growth models to generate short-term (i.e., 30-day ahead) forecasts for Colombia. These models have been applied to various infectious diseases including SARS, foot and mouth disease, Ebola [47–49] and the current COVID-19 outbreak [50–52]. The phenomenological growth models applied in this study include the two models defined by one scalar differential equation namely the generalized logistic growth model (GLM) [48] and the Richards growth model [53]. Additionally we apply the sub-epidemic wave model [47] which captures diverse epidemic trajectories such as the multimodal outbreaks. The forecasts obtained from these dynamic growth models can assess the potential scope of the pandemic in near real-time, provide insights on the contribution of disease transmission pathways, predict the impact of control interventions and evaluate optimal resource allocation to inform public health policies. A detailed description of these models is provided in the supplemental file.

### Model calibration and forecasting approach

We conducted three 30-day ahead short-term forecasts for each model. We utilize the national and regional level data retrieved from the Colombian Ministry of Health according to the dates of symptom onset as of June 30, 2021. Each forecast was fitted to the daily case counts by dates of symptom onset between April 1, 2021 and May 30, 2021 (60 days calibration period). We drop the last 30 days of data to assess the performance of our 30-day ahead short-term forecasts. Details of the model calibration and forecasting approach are provided in the supplemental file.

### Performance metrics

We utilize the following four performance metrics to assess the quality of our model fit and the 30-day ahead short-term forecasts: the mean absolute error (MAE) [54], the root mean squared error (RMSE) [55], the coverage of the 95% prediction intervals (95% PI) [55], and the mean interval score (MIS) [55] for each of the three models: GLM, Richards model and the sub-epidemic wave model. A detailed description of the performance metrics is provided in the supplemental file.

### Reproduction number

We estimate the effective reproduction number, *R_t_*, for the early ascending phase of the COVID-19 pandemic in Colombia and the time dependent instantaneous reproduction number *R_t_* throughout the pandemic for the national and regional COVID-19 curves. The reproduction number, *R_t_*, is the key parameter that characterizes the average number of secondary cases generated by a primary case at calendar time *t* during the outbreak. This quantity is crucial for identifying the magnitude of public health interventions required to contain an epidemic [56–58]. The estimates of *R_t_* indicate whether widespread disease transmission continues (*R_t_* >1) or disease transmission declines (*R_t_* <1). Therefore, to contain an outbreak, it is vital to maintain *R_t_* <1.

### Reproduction number *R_t_*, using the generalized growth model (GGM)

We estimate the national and regional reproduction numbers by calibrating the GGM to the early growth phase of the pandemic [59]. The description of the GGM is provided in the supplemental file. The generation interval of SARS-CoV-2 is modeled with assumption of a gamma distribution with a mean of 5.2 days and a standard deviation of 1.72 days [60]. We first characterize the daily incidence of local cases using the GGM after adjusting for imported cases. We estimate the growth rate parameter *r*, and the deceleration of growth parameter, *p* from the GGM. The GGM model is used to simulate the progression of local incidence cases *I_i_* and imported case incidence *J_i_* at calendar time *t_i_*. This is followed by the application of the discretized probability distribution of the generation interval, denoted by *ρ_i_*, to the renewal equation to estimate the reproduction number at time *t_i_* [61–63]:

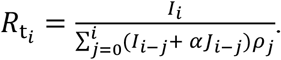

The factor *J_i_* represents the imported cases at time *t_i_*, *I_i_* denotes the local case incidence at calendar time *t_i_* and *ρ_i_*. represents the discretized probability distribution of the generation interval. The factor 0 ≤ *α* ≤ 1 represents the relative contribution of imported cases to secondary disease transmission. We perform sensitivity analysis by setting *α* =0.15 and *α* =1.00 [64]. The numerator represents the total new cases *I_i_*, and the denominator represents the total number of primary cases that contributes to the generation the new cases *I_i_* (i.e., as secondary cases) at time *t_i_*. Hence, *R_t_*, represents the average number of secondary cases generated by a single case at calendar time *t*. The uncertainty bounds around the curve of *R_t_* are derived directly from the uncertainty associated with the parameter estimates (*r*, *p*) obtained from the GGM. We estimate *R_t_* for 300 simulated curves assuming a negative binomial error structure where the variance is assumed to be three times the mean based on the noise of the data [65].

Since the national and regional epidemic curves have a distinct date of onset according to the seeding of the first local case, therefore, *R_t_* for the national and regional level is estimated starting on the date of onset of the first local case. For the national and regional epidemic curves, we estimate *R_t_* for the first 30 days (Table 1) .

**Table 1:**
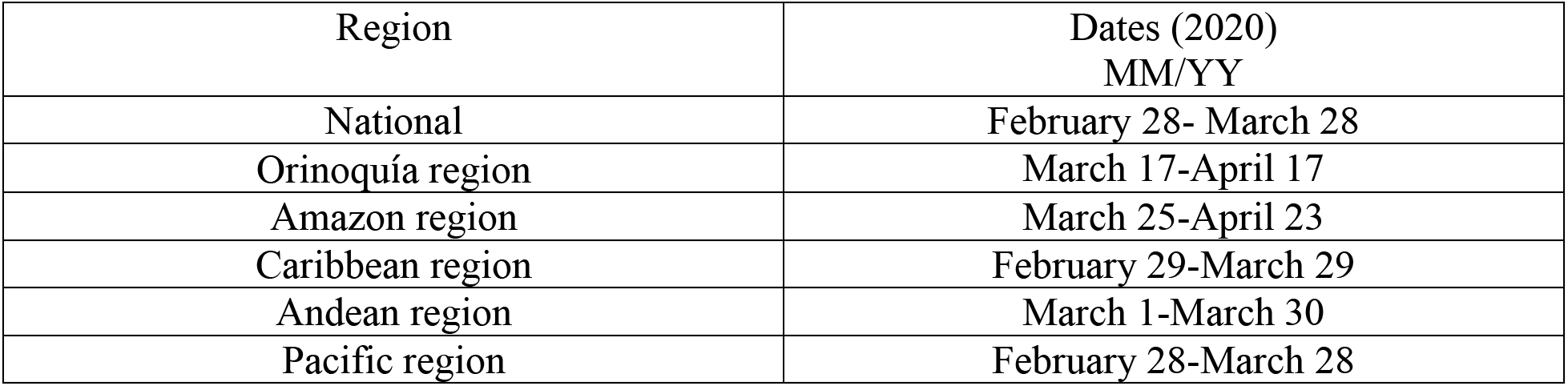
Dates for *R_t_* estimation for the national and regional epidemic curves for the first 30 epidemic days.

| Region | Dates (2020)<br>MM/YY |
| --- | --- |
| National | February 28- March 28 |
| Orinoquía region | March 17-April 17 |
| Amazon region | March 25-April 23 |
| Caribbean region | February 29-March 29 |
| Andean region | March 1-March 30 |
| Pacific region | February 28-March 28 |

### Instantaneous reproduction number *R_t_*, using the Cori method

We estimated the regional and national instantaneous reproduction numbers using the local and imported case incidence data by the symptoms onset date as of May 30, 2021. The total number of incident cases arising at time step t, *I_t_*, is the sum of the number of local incident cases 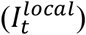 and imported incident cases 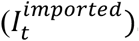 Hence,

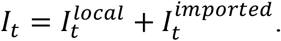

We assume that if the imported cases exist, they can be distinguished from the local cases through epidemiological investigations so that 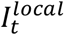 and 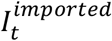 can be observed at each time step [63, 66].

The time dependent (instantaneous) *R_t_* is defined as the ratio of the number of new locally infected cases generated at calendar time *t* 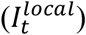 and the total infectiousness across all infected individuals at time *t* given by 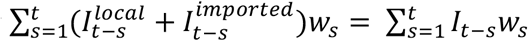 [67, 68] . Hence *R_t_* can be written as

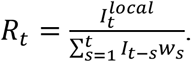

In this equation, the factor *w_s_* represents the infectivity function, which is the infectivity profile of the infected individual. The infectivity function is dependent on the time since infection (*s)*, but is independent of the calendar time (*t*) [69, 70].

The term 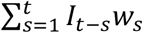 describes the sum of infection incidence up to time step *t* − 1, weighted by the values of infectivity function *w_s_*. The distribution of the generation time can be applied to approximate *w_s_*, however, since the time of infection is a rarely observed event, measuring the distribution of generation time becomes difficult [67]. Therefore, the time of symptom onset is usually used to estimate the distribution of the serial interval (SI), which is defined as the time interval between the dates of symptom onset among two successive cases in a disease transmission chain [71].

The infectiousness of a case is a function of the time since infection, which is proportional to *w_s_* if the timing of infection in the primary case is set as time zero of *w_s_* and we assume that the generation interval equals the serial interval (SI). The SI was assumed to follow a gamma distribution with a mean of 5.2 days and a standard deviation of 1.72 days [60]. Analytical estimates of *R_t_* were obtained within a Bayesian framework using the EpiEstim R package in R version 4.0.3. [71]. The values for *R_t_* were estimated using 1-weekly sliding window. We reported the median and 95% credible interval (CrI).

### Reproduction number, *R,* from the genomic analysis

In order to estimate the early reproduction number for the SARS-CoV-2 between February 27, 2020 and April 5, 2020 from the genomic data, 136 SARS-CoV-2 genomes from Colombia and their sampling times were were obtained from the GISAID (global initiative on sharing all influenza data) repository [72]. Short sequences and sequences with significant number of gaps and non-identified nucleotides were removed. For clustering, they were complemented by sequences from other geographical regions across the globe. We used the sequence subsample from Nextstrain (www.nextstrain.org) global analysis as of July 4, 2021. These sequences were aligned to the reference genome obtained from the literature [73] using Muscle [74] and trimmed using Mega 7 (Molecular Evolutionary Genetics Analysis version: 7) [75].

The largest Colombian cluster that possibly corresponds to within-country transmissions has been identified using hierarchical clustering of sequences. Phylodynamics analysis of that cluster was carried out using BEAST version 2 (Bayesian Evolutionary Analysis by sampling trees) [76]. We used a strict molecular clock and a tree prior with exponential growth coalescent. Markov Chain Monte Carlo (MCMC) sampling was run for 10,000,000 iterations, and the parameters were sampled every 1000 iterations. The exponential growth rate *f* estimated by BEAST was used to calculate the basic reproduction number, *R*_0_ for the early part of the epidemic. We utilized the standard assumption that SARS-CoV-2 generation intervals (times between infection and onward transmission) are gamma-distributed [77]. In this case, *R*_0_ can be estimated as 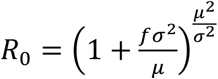. where *μ* and *σ* are the mean and standard deviation of the gamma distribution, respectively. The values of *μ* = 5.2 days and *σ* = 1.72 days were taken from the study [60].

### Spatial analysis

We quantify and statistically analyze the shapes of the incidence rate curves. Shape analysis of the national and regional incidence rate curves was performed following the analytical methodology as described in previous literature [78] including steps to pre-process the daily cumulative COVID-19 case data based on the dates of symptom onset. Then, we analyze the shapes of these rate curves to compare, cluster and summarize incidence rates. The pre-processing steps applied to the COVID-19 daily count data are as follows:

a. Smoothing: Cumulative case curves are smoothed over 10 days using the smooth function in MATLAB.
b. Time differencing: If *f_i_*(*t*) denotes the given cumulative number of confirmed cases for department *i* on day *t*, then the growth rate per day at time *t* is given by *g_i_*(*t*) = *f_i_*(*t*) – *<u>f_i_</u>*(*t*–1).
c. Rescaling: Each curve is rescaled by dividing each *g_i_* by the total number of confirmed cases.
d. Smoothing: We then smoothed the normalized curves again over a 5-day span using the smooth function in MATLAB.

This process is shown in Fig. 9. To identify the clusters by comparing the curves, we used a simple metric. For any two rate curves, *h_i_* and *h_j_*, we compute the norm *||h_i_ −h_j_||,* where the double bars denote the *L^2^* norm of the difference function, that is,

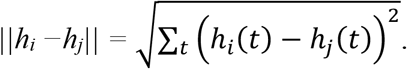

To perform clustering of thirty six curves into smaller groups, we apply the dendrogram function in MATLAB using the “ward” linkage as explained in reference [79]. The number of clusters is determined empirically based on the display of overall clustering results. After clustering the departments and administrative divisions into different groups, we derived the average curve for each cluster after using a time wrapping algorithm as performed in previous works [79, 80].

### Twitter data analysis

To observe any relationship between the COVID-19 cases by date of symptom onset and the frequency of tweets indicating stay-at-home orders we used a public dataset of 270 million tweets of COVID-19 chatter [46]. The frequency of tweets indicating stay-at-home order is used to infer people’s compliance with the stay-at-home-orders implemented to avoid spread of the virus by practicing social distancing. Tweets indicate the number of the people in favor of lockdowns and show how these numbers have varied throughout the pandemic. We removed all retweets and tweets not in the Spanish language to get to the plotted data. We also filtered by the following hashtags: #quedateencasa (stay-at-home), and #trabajardesdecasa (work-from-home), which are two of the most used hashtags when users refer to the COVID-19 pandemic and their engagement with health measures. Lastly, our analysis was restricted to the tweets that originated from Colombia. A set of 393,165 unique tweets were gathered from March 12, 2020, to June 30, 2021. The usage of Twitter in Colombia is less compared to other Latin American countries like Mexico and Chile [81], hence we observe a lower proportion of filtered tweets. We overlay the curve of filtered tweets and the epidemic curve in Colombia to observe any relation between the shape of the epidemic trajectory and the shape of curve for the frequency of filtered tweets during the established time-period. The correlation coefficient between the daily cases by dates of onset and frequency of filtered tweets is also estimated.

### Mobility data analysis

We utilize the R code developed by Healy [82] to analyze the time series data for Colombia from March 6, 2020 to June 30, 2021, for two modes of mobility: driving and walking derived from Apple’s mobility reports. We analyzed the mobility trends to look for any commonality in pattern with the epidemic curve of COVID-19. The time series for mobility requests is decomposed into trends, weekly and remainder components. The trend is fitted by locally weighted regression fitted to the data and the remainder is any residual leftover on any given day after accounting for the underlying trend and normal daily fluctuations.

To analyze the time series for Colombia from March 6, 2020, to June 30, 2021, we analyzed the Google mobility trends for visits to retail and recreation, grocery and pharmacy, parks, transit stations, workplaces, and residential areas compared to the baseline which is the median value, for the corresponding day of the week, during the 5 weeks from January 3, 2020 to February 6, 2020. For each region category, the baseline is not a single value; it comprises of seven individual values. The same number of visitors on two different days of the week results in different percentage changes. The analysis was performed using R software, version 4.0.3.

## RESULTS

A timeline showing the major events during the COVID-19 pandemic in Colombia is presented in Fig 1. As of June 30, 2021, Colombia has reported 3,763,727 cases based on the dates of symptom onset. The highest number of cases have been reported in the Andean region (2,429,604 cases) followed by the Caribbean region (754,835 cases), Pacific region (418,725 cases), Orinoquía region (113,500 cases) and the Amazon region (47,063 cases). The COVID-19 epidemic curve in Colombia shows a five-modal pattern with the first peak occurring in mid-July 2020 after the phased reopening of the country. It was followed by a second peak occurring in mid-October 2020 after the implementation of selective isolation and responsible social distancing interventions. The third peak occurred at the beginning of January 2021 and the fourth peak of COVID-19 cases can be observed in the beginning of April 2021 (Fig 2.). A most recent surge in cases can be observed as of May 2021. This most recent uptick in cases has led to the selective curfews in cities in the country based on the ICU occupancy rates in the hospitals.

**Fig 1.**
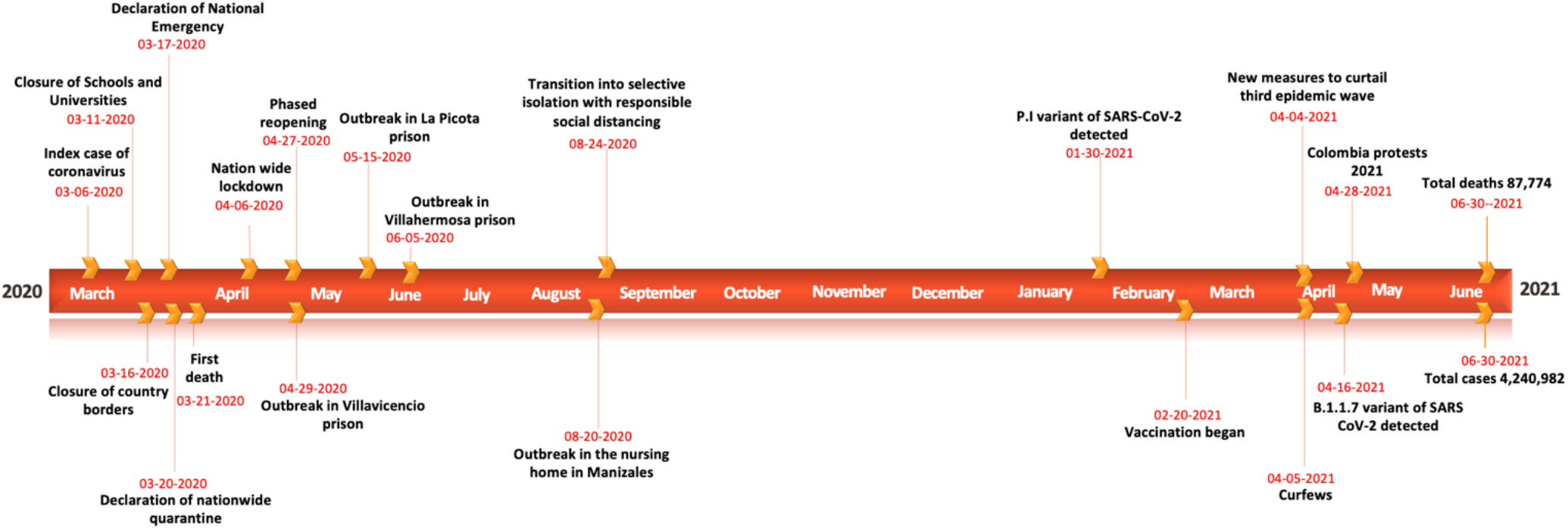
Timeline of the COVID-19 pandemic in Colombia as of June 30, 2021.

**Fig 2.**
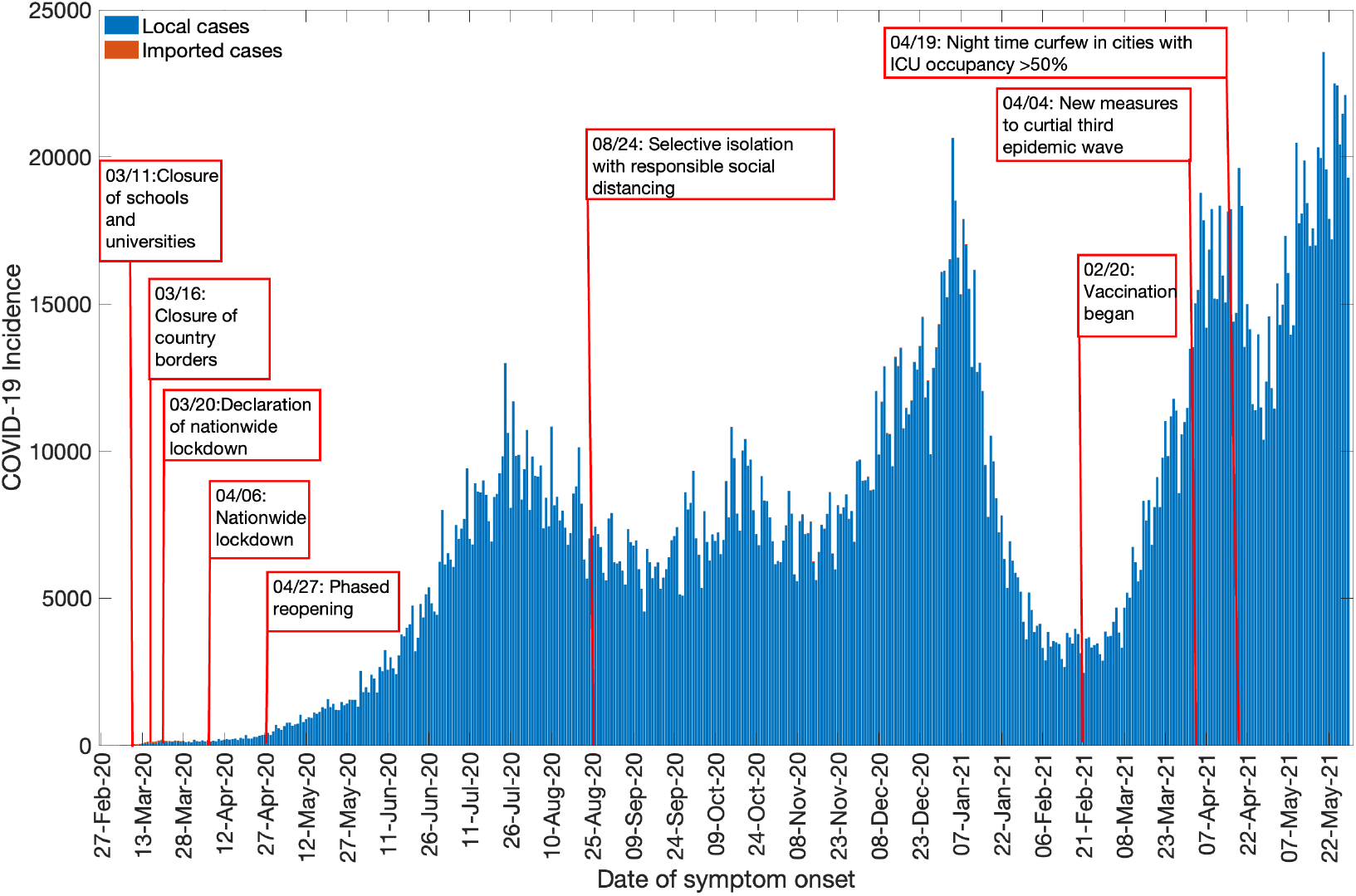
Epidemic curve of the COVID-19 pandemic in Colombia as of May 30, 2021

### Model calibration and forecasting performance

We compare the results from model calibration and 30-day ahead forecasting across three models: GLM, Richards and the sub-epidemic wave model. We use the last 60 days of the epidemic data according to the dates of symptom onset for model calibration. Model calibration shows that at the regional and national level, the sub-epidemic model outperformed the GLM and Richards growth model in terms of the three performance metrics (MAE, MIS and 95% PI coverage), whereas GLM performed better than the Richards and sub-epidemic wave model in terms of RMSE (Table 2). Therefore, the sub-epidemic wave model can be declared a winner for the calibration period. The model fits exhibited sub-exponential growth dynamics for three models (*p*∼0.6-0.8) at the national and regional levels. The calibration performances for each region and the national data are listed in Table 2.

**Table 2:**
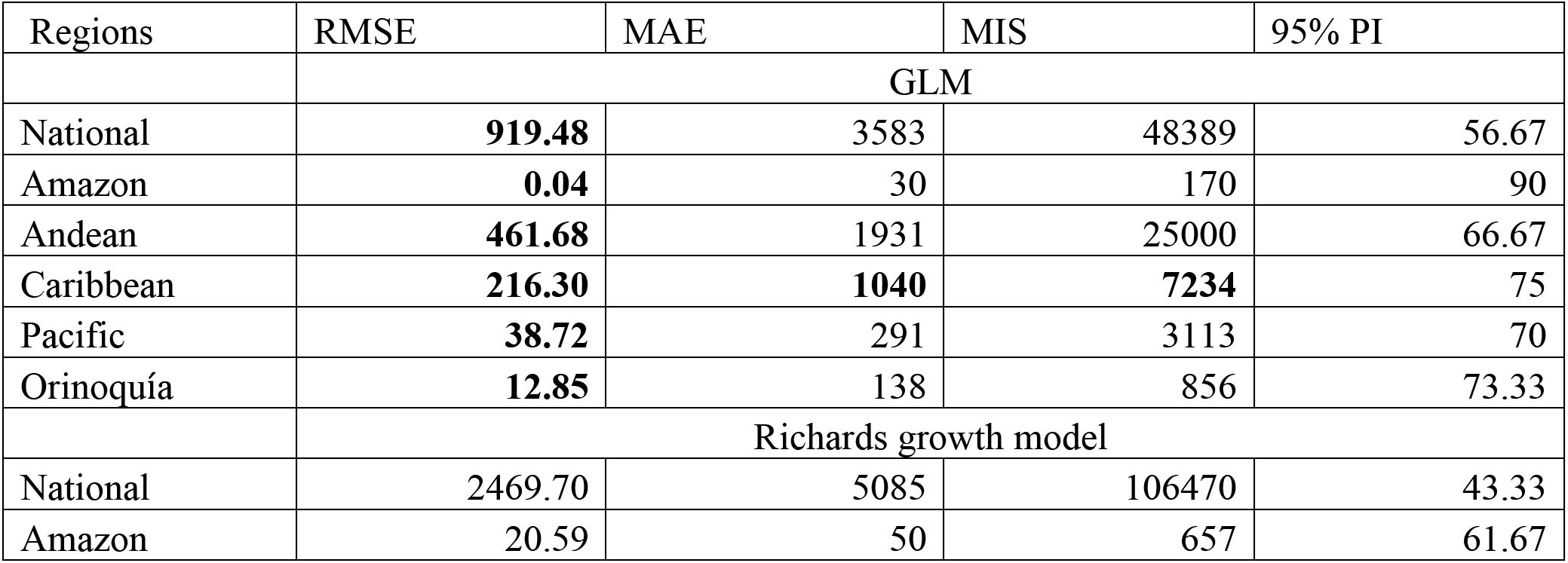
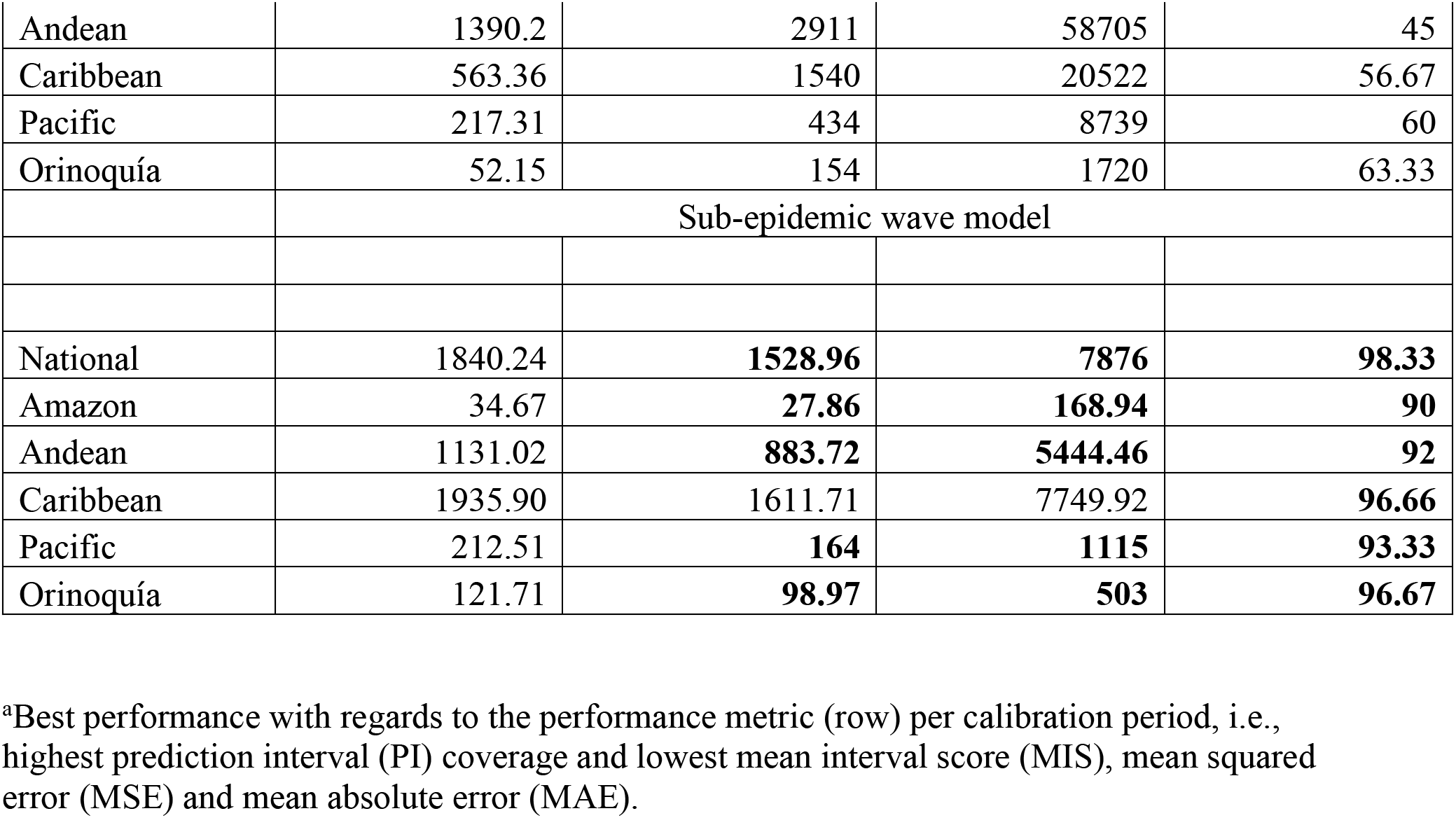
Comparison of model performance metrics by calibrating the GLM, Richards and the sub-epidemic model for the most recent 60 epidemic days (04/01/2021-05/30/2021) at the national and regional level. Higher 95% PI coverage and lower RMSE, MAE and MIS represent better performance.

In terms of the forecasting performance, the sub-epidemic wave model performed better than the GLM and Richards model for the national, Pacific and the Orinoquía region. The GLM performed better than the other two models for the Caribbean region and the Richards model performed better than the other two models for the Andean and Amazon region (Table 3).

**Table 3:**
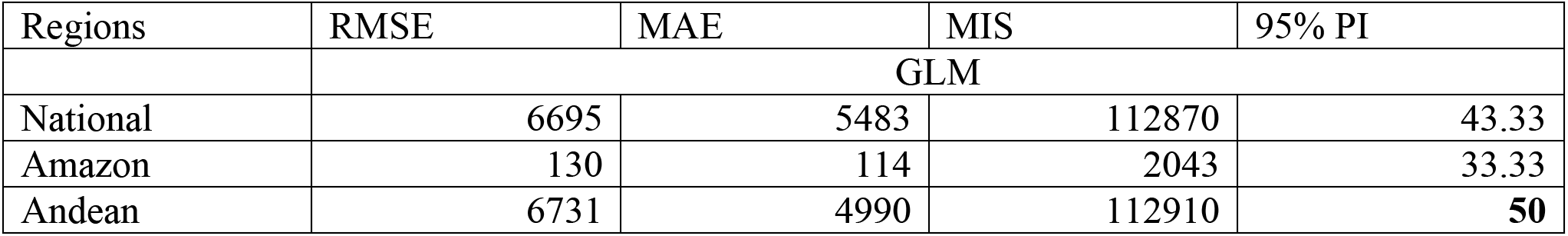
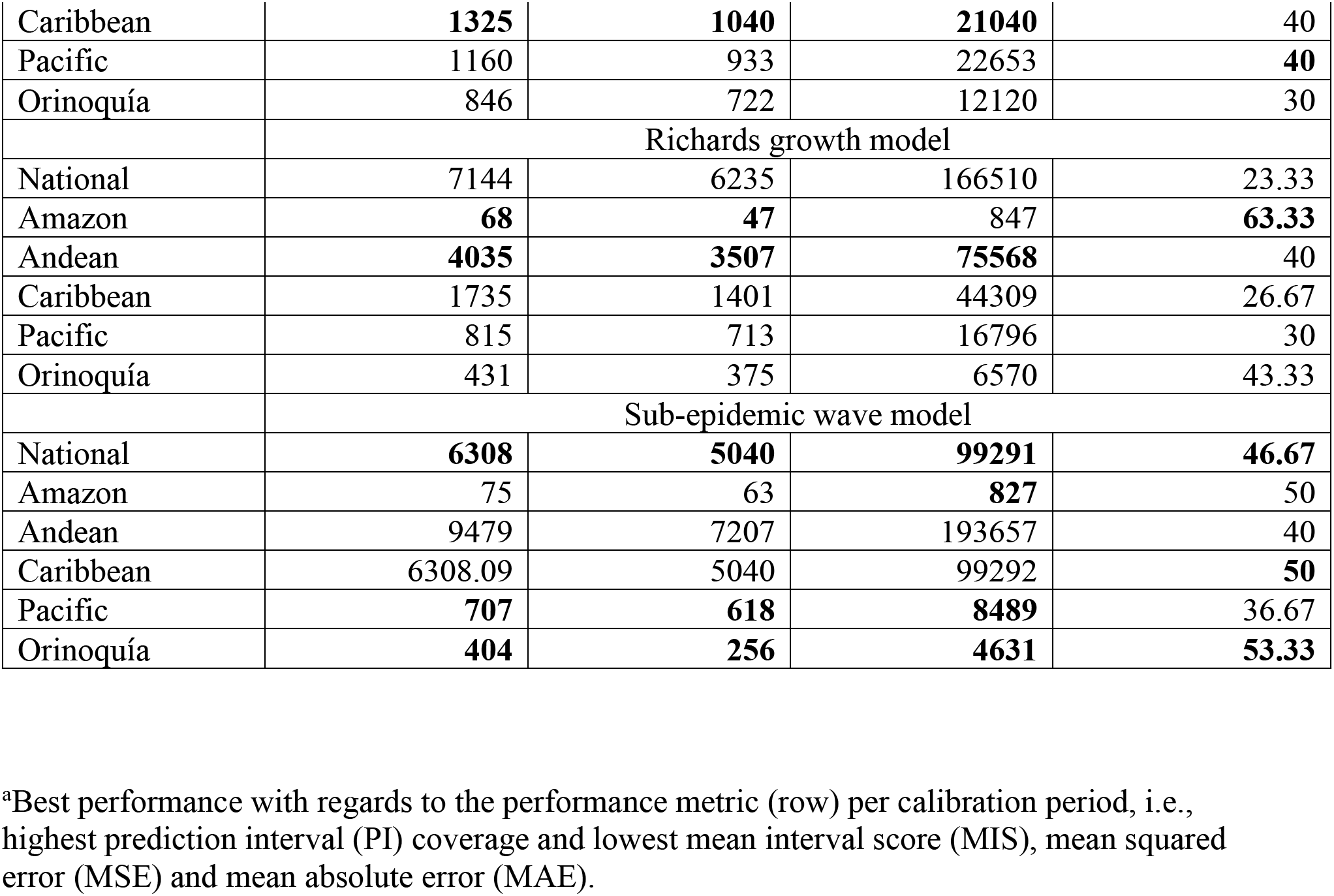
Comparison of 30-day ahead forecasting performance (061/2021-06/30/2021) by calibrating the GLM, Richards and the sub-epidemic model for the most recent 60 epidemic days (04/01/2021-05/30/2021) at the national and regional level. Higher 95% PI coverage and lower RMSE, MAE and MIS represent better performance.

### 30-day ahead forecasts

Calibrating our models from April 01, 2021 to May 30, 2021 and generating the 30-day ahead forecasts (between June 01, 2021 and June 30, 2021) utilizing the GLM indicates a downwards trend for the national and Caribbean region, whereas an upward trend in case incidence for the Pacific, Andean, Amazon and Orinoquía region (Fig 3). The Richards model however indicates a downward trend in the cases by dates of symptom onset for all the national and regional level forecasts (Fig 4). On the other hand, the sub-epidemic model captures the multiple sub-epidemics comprising the coarse COVID-19 epidemic wave of Colombia. The sub-epidemic model predicts an upward trend in cases for the national and Andean region suggesting an increase in case incidence. Whereas, for the Pacific and Orinoquía region, the sub-epidemic model predicts a downward trend with a stable case incidence pattern predicted for the Amazon and Caribbean regions. According to the GLM, the COVID-19 pandemic in Colombia can accumulate on average ∼464,174 cases (95% PI: 337,863-611,960) at the national level in the next 30 days (between June 01, 2021 and June 30, 2021). Similarly, the Richards model indicates that the COVID-19 epidemic could accumulate on average ∼294,683 cases (95% PI: 207,786-402, 811) and the sub-epidemic wave model predicts an accumulation of 693,257 (95% PI: 541,353-884,885) cases at the national level in the next thirty days.

**Fig 3.**
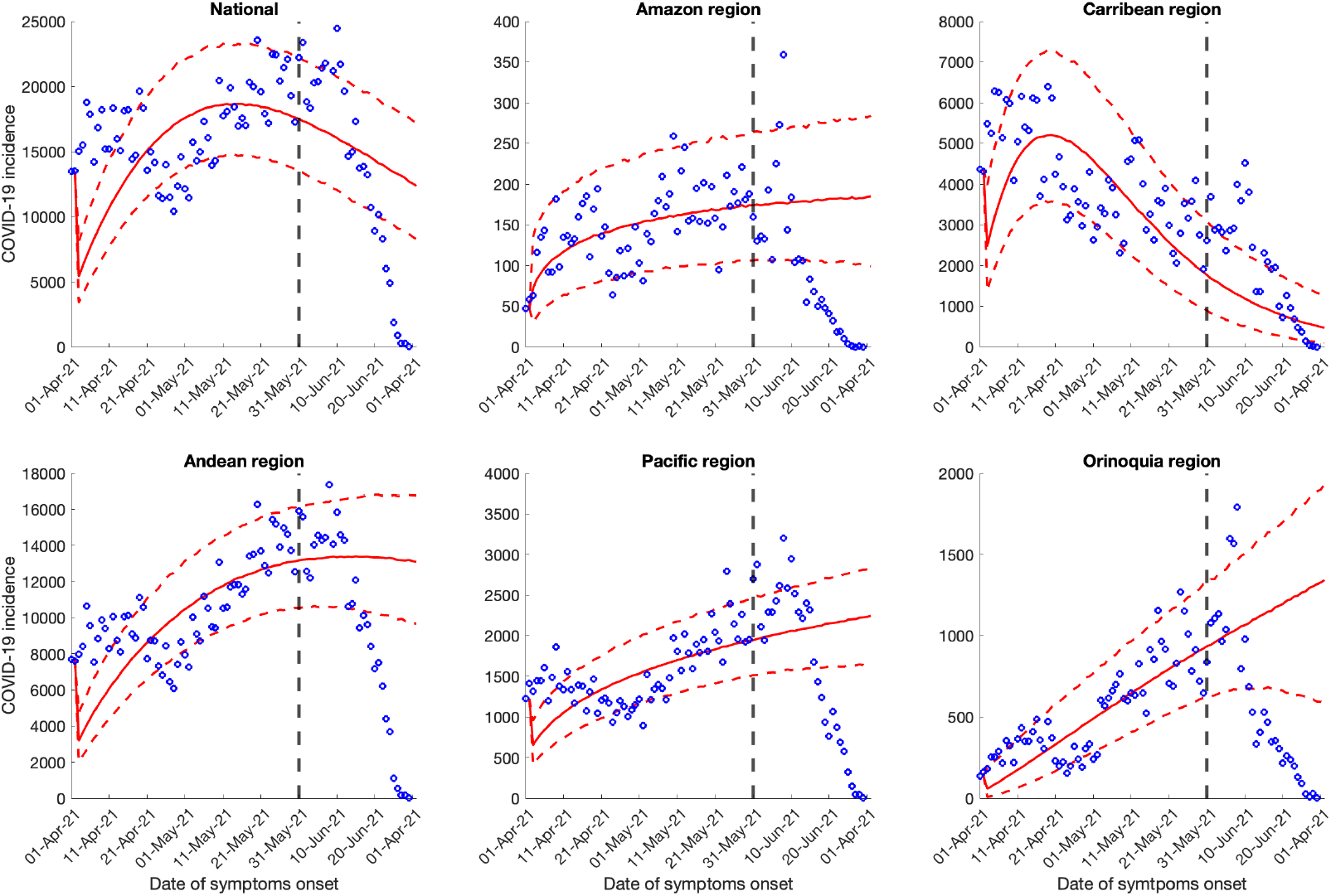
30-days ahead forecasts of the national and regional COVID-19 epidemic curves in Colombia by calibrating the GLM model from April 1, 2021 to May 30, 2021. Blue circles correspond to the data points; the solid red line indicates the best model fit, and the red dashed lines represent the 95% prediction interval. The vertical black dashed line represents the time of the start of the forecast period.

**Fig 4.**
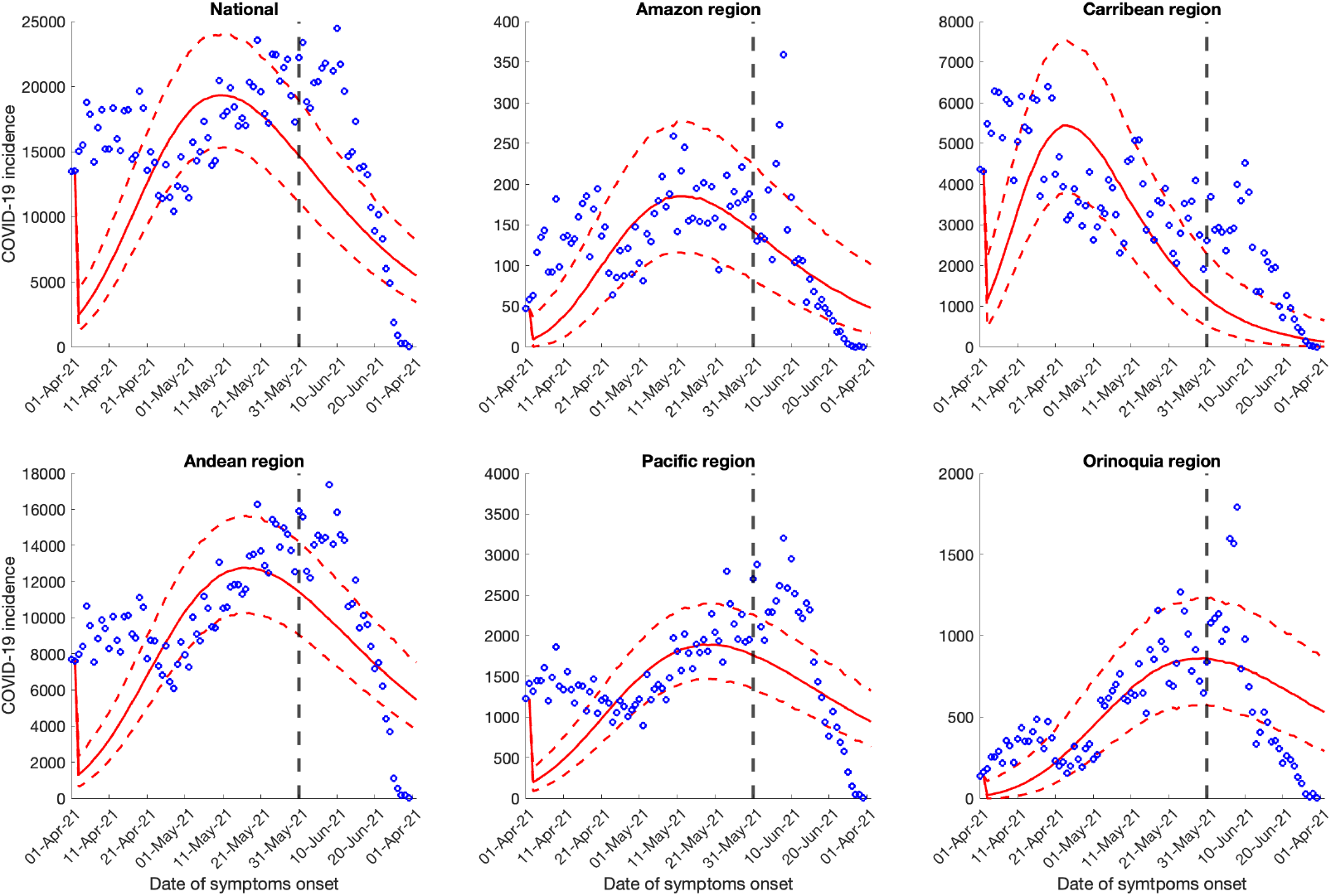
30-days ahead forecast of the national and regional COVID-19 epidemic curves in Colombia by calibrating the Richards model from April 1, 2021 to May 30, 2021 region. Blue circles correspond to the data points; the solid red line indicates the best model fit, and the red dashed lines represent the 95% prediction interval. The vertical black dashed line represents the time of the start of the forecast period.

At the regional level, the three models predict on average an additional 2749-5568 cases in the next 30 days for the Amazon region. The upper bounds of the 95% PI of GLM and sub-epidemic model suggest on average an additional 8497 maximum cases during the forecast period. The average 30-day ahead forecast for the Caribbean region from sub-epidemic wave model predicts ∼98,219 additional cases. The GLM model however predicts an additional case count of ∼30,280 cases and the Richards model further under-predicts the case counts, forecasting an average of ∼15,201 cases in the next thirty days. The upper bounds of the 95% PI for the GLM, Richards and sub-epidemic wave model ranges from 30,280-188,895 cases. For the Andean region the three models predict on average between 257,321-553,262 cases in the next thirty days. The upper bounds of the 95% PI range from 333,143-673,006 additional cases. The average 30-day ahead forecasts for the Pacific region from the Richards and sub-epidemic model predict an addition of 41,780 and 53,381 cases respectively in the next thirty days with the upper bounds of the 95% PI ranging from 55,523-69,360 cases. However, the GLM model has significantly higher upper bounds, suggesting a possibility to accumulate up to an average of 82,344 cases in the next thirty days. Lastly, for the Orinoquía region the upper bounds of the 95% PI for the three models ranges from 18,162-50,471 cases in the next thirty days with an average of 11,017 additional cases as predicted by the sub-epidemic wave model compared to the additional 35,511 and 22,198 cases predicted by the GLM and Richards model, respectively (Figs 3, 4 and 5).

**Fig 5.**
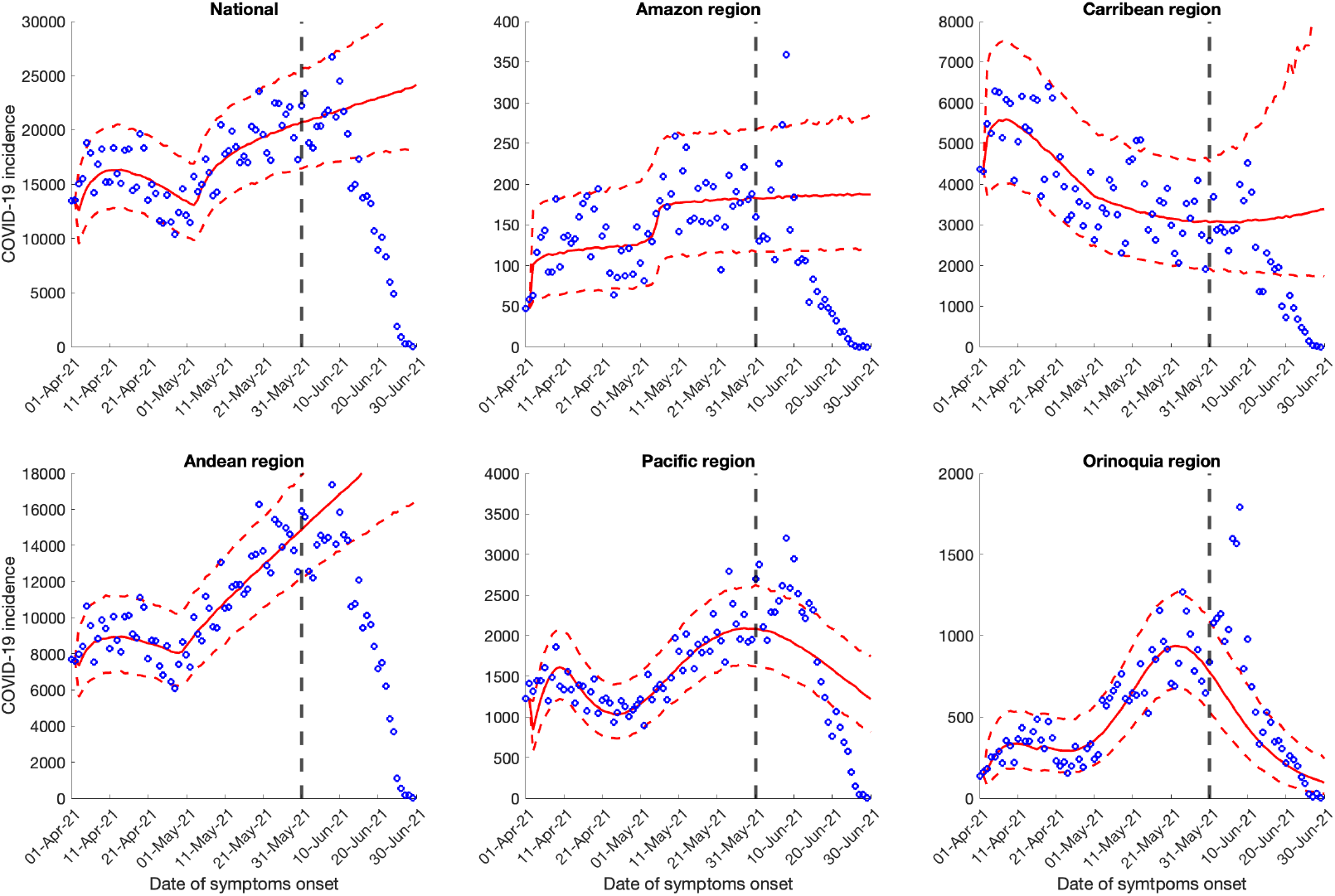
30-days ahead forecast of the national and regional COVID-19 epidemic curves in Colombia by calibrating the sub-epidemic wave model from April 1, 2021 to May 30, 2021 region. Blue circles correspond to the data points; the solid reds line indicates the best model fit, and the red dashed lines represent the 95% prediction interval. The vertical black dashed line represents the time of the start of the forecast period.

### Reproduction number

#### Estimate of reproduction number, *R_t_* from case incidence data

The reproduction number for the early ascending growth phase of the epidemic from the case incidence data (February 27, 2020 to March 27, 2020) using GGM was estimated at *R_t_* ∼1.30 (95% CI:1.20-1.50) at *α* = 0.15 for the national data. The growth rate parameter, *r*, was estimated at 1.40 (95% CI: 0.91-2.0) and the deceleration of growth parameter, *p*, was estimated at 0.64 (95% CI: 0.56-0.71), indicating early sub-exponential growth dynamics of the epidemic (Fig 6). Simultaneously, the estimates of *R_t_* for all the regions remained consistently above 1.20 (between ∼1.20-2.22) for the early ascending phase of the pandemic with the Amazon region showing the highest estimate of reproduction number, *R_t_* ∼2.2 followed by Orinoquía region with *R_t_* ∼1.8. The estimates of *R_t_* for the Andean, Pacific and Caribbean regions remained between *R_t_* ∼1.2-1.4. All regions except the Amazon region depict sub-exponential growth dynamics for the COVID-19 pandemic in Colombia with the deceleration of growth parameter, *p*, estimated between 0.54-0.86. The Amazon region shows almost exponential growth dynamics with the deceleration of growth parameter, *p* ∼0.95 (95% CI: 0.74, 1.00). In contrast, Andean and Caribbean regions show an almost linear pattern of the epidemic trajectory with the deceleration of growth parameter, *p* ∼0.58 (95% CI:0.48-0.69) and *p* ∼0.59 (95% CI: 0.34, 0.87) respectively. The sensitivity analysis shows that the estimates of reproduction numbers do not vary significantly at *α* = 0.15 and *α* = 1.00 (Tables 4 and 5). Fig 6. Upper panel: Reproduction number for Colombia with 95% CI estimated using the GGM model. The estimated reproduction number of the COVID-19 epidemic in Colombia as of March 27, 2020, is 1.30 (95% CI: 1.20, 1.50). The growth rate parameter, r, is estimated at 1.40 (95%CI: 0.91, 2.0) and the deceleration of growth parameter, p, is estimated at 0.64 (95%CI: 0.56, 0.71) at *α* = 0.15.

**Fig 6.**
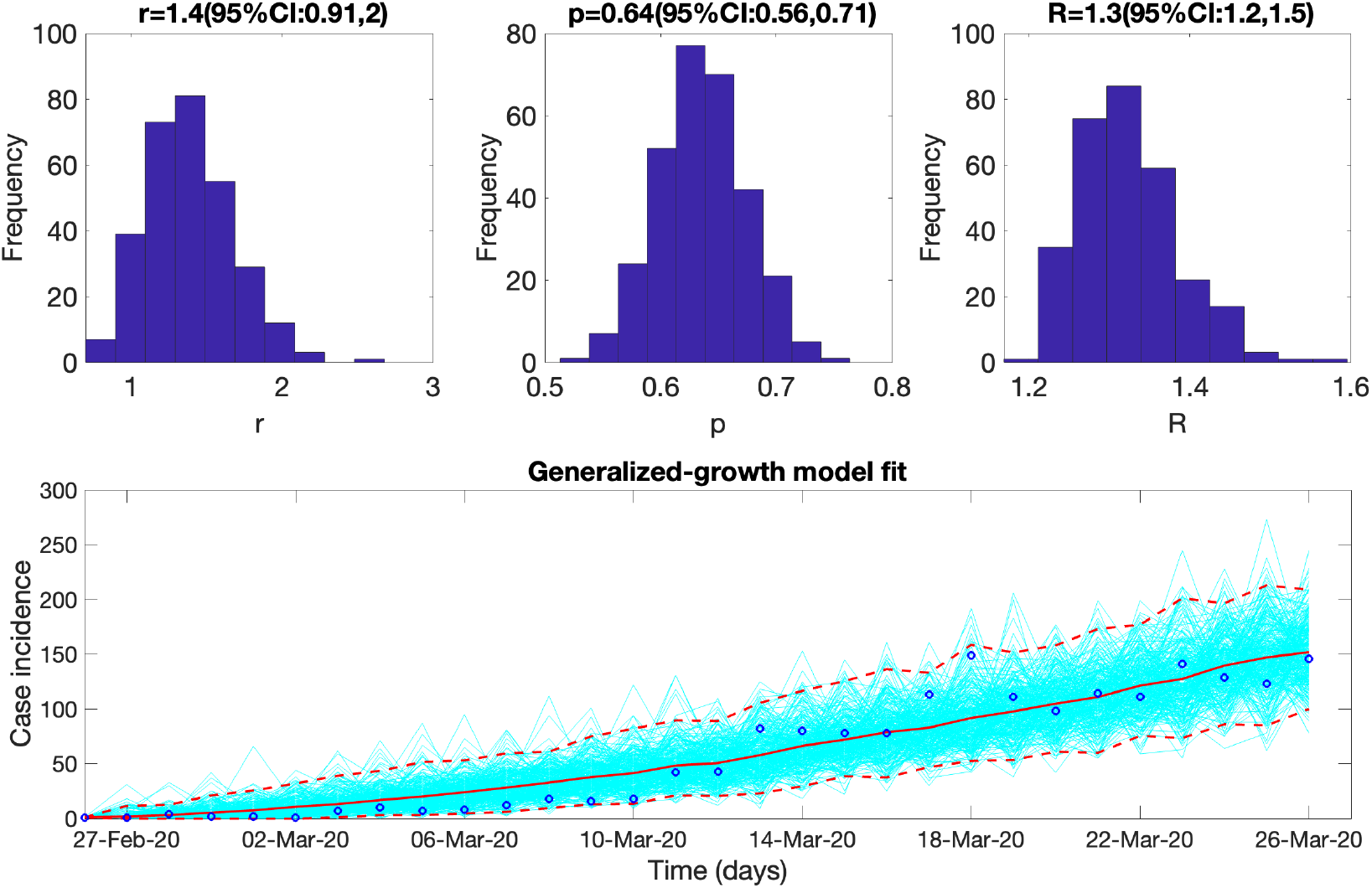
Upper panel: Reproduction number for Colombia with 95% CI estimated using the GGM model. The estimated reproduction number of the COVID-19 epidemic in Colombia as of March 27, 2020, is 1.3 (95% CI: 1.2, 1.4). The growth rate parameter, *r*, is estimated at 1.4 (95%CI: 0.9, 1.2) and the deceleration of growth parameter, *p*, is estimated at 0.64 (95%CI:0.56, 0.71) at *α* = 0.15. Lower panel: The lower panel shows the GGM fit to the case incidence data for the first 30 days (February 27- March 27, 2020).

**Table 4:** Estimates of reproduction number (*R)*, growth rate parameter *(r)* and the deceleration of growth parameter (*p*) obtained from the renewal equation method utilizing the GGM for the early ascending phase of the epidemic (30 days) at the national and regional level at *α* = 0.15.

| Region | $r$ | $p$ | $R$ |
| --- | --- | --- | --- |
| National | 1.4 (95% CI: 0.91,2.0) | 0.64 (95% CI: 0.56, 0.71) | 1.3 (95% CI: 1.2,1.5) |
| Amazon | 0.23 (95% CI: 0.18, 0.4) | 0.95 (95% CI: 0.74, 1.00) | 2.2 (95% CI:1.5, 2.6) |
| Andean | 1.7 (95% CI:0.88, 2.8) | 0.58 (95% CI:0.48, 0.69) | 1.2 (95% CI: 1.1, 1.4) |
| Caribbean | 0.85 (95% CI:0.27, 1.9) | 0.59 (95% CI:0.34, 0.87) | 1.3 (95% CI:1.1,1.8) |
| Pacific | 0.63 (95% CI:0.25, 1.3) | 0.69 (95% CI:0.48, 0.9) | 1.4 (95% CI:1.1, 2.0) |
| Orinoquía | 0.25 (95% CI:0.16, 0.59) | 0.87 (95% CI: 0.58, 1.00) | 1.8 (95% CI:1.3, 2.4) |

**Table 5:** Estimates of reproduction number (*R*), growth rate parameter *(r*) and the deceleration of growth parameter (*p*) obtained from the renewal equation method utilizing the GGM for the early ascending phase of the epidemic (30 days) at the national and regional level at *α* = 1.00.

| | $r$ | $p$ | $R$ |
| --- | --- | --- | --- |
| National | 1.4 (95% CI: 0.87 2.1) | 0.64 (95% CI: 0.56, 0.72) | 1.1 (95% CI:0.97, 1.2) |
| Amazon | 0.23 (95% CI:0.18, 0.39) | 0.94 (95% CI:0.77, 1.0) | 2.1 (95% CI:1.4, 2.5) |
| Andean | 1.7 (95% CI:1.0, 2.8) | 0.58 (95% CI: 0.48, 0.67) | 1.0 (95% CI:0.94, 1.2) |
| Caribbean | 0.82 (95% CI:0.19, 2.00) | 0.61 (95% CI:0.34, 0.99) | 1.3 (95% CI:1.0, 2.3) |
| Pacific | 0.61( 95% CI:0.23, 1.2) | 0.69 (95% CI:0.51, 0.95) | 1.2 (95% CI:0.93, 1.9) |
| Orinoquía | 0.26 (95% CI: 0.16, 0.53) | 0.86 (95% CI:0.6, 1.0) | 1.8 (95% CI:1.3, 2.4) |

### Estimate of instantaneous reproduction number, *R_t_*

The early estimate of instantaneous reproduction number at the national level for the first 30 epidemic days exhibits a steep decline. The reproduction number was estimated at, *R_t_* ∼2.85 (95% CrI: 1.91-4.09) on March 6, 2020, which declined to *R_t_* ∼0.81 (95% CrI : 0.76-0.85) by March 27, 2020 (Figs 7 and 8). At the national level the instantaneous reproduction number has remained consistently above *R_t_* ∼1.00 between April and July 3, 2020, after which the reproduction number declined to less than 1.00 (*R_t_* ∼0.9) until September 22, 2020. Since the end of September 2020, *R_t_* has fluctuated around ∼1.00 (*R_t_* ∼ 0.8-1.4) with the most recent estimate of *R_t_* ∼1.03 (95% CrI: 1.02-1.03) as of May 30, 2021 (Fig 8).

**Fig 7.**
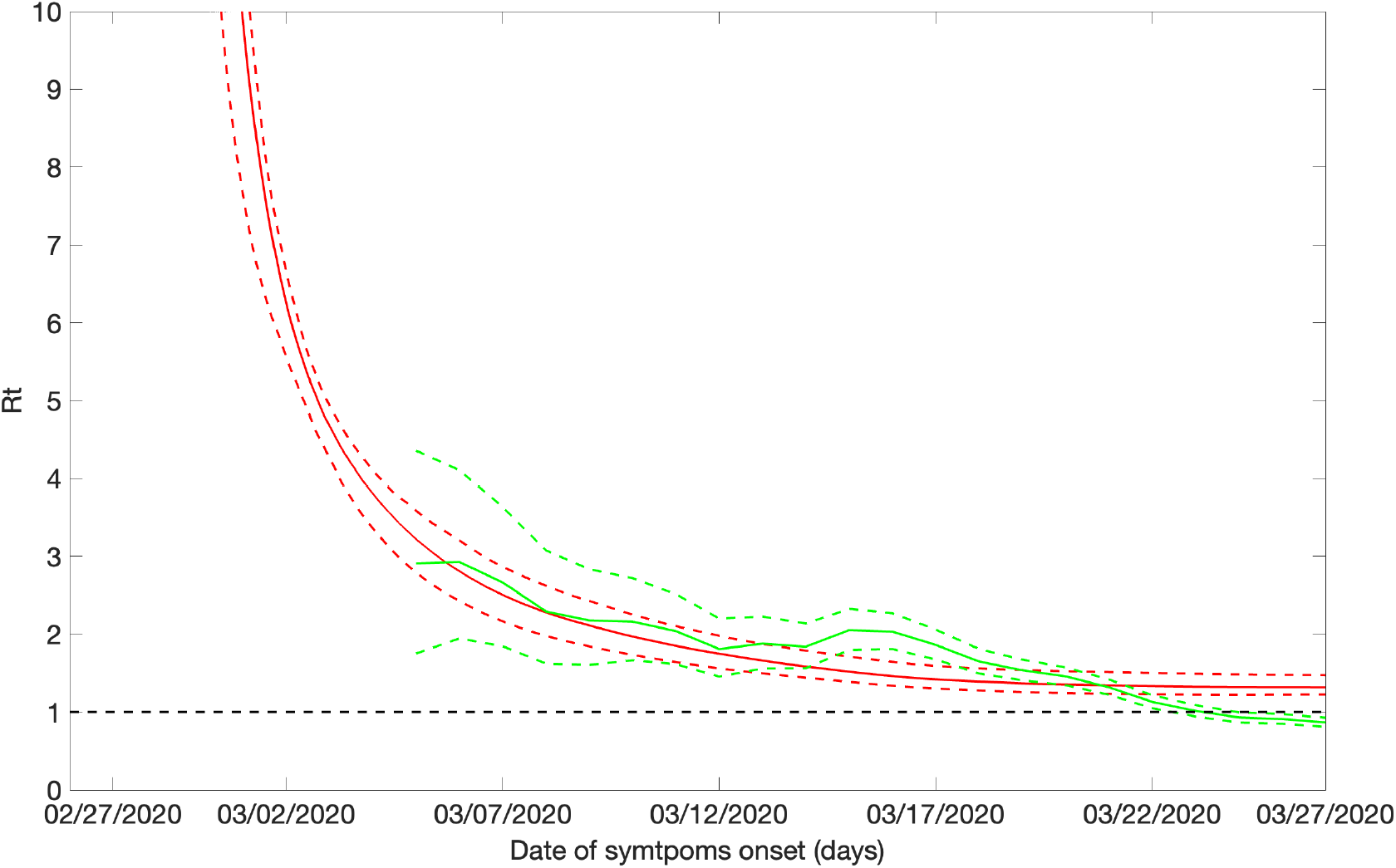
Instantaneous reproduction number with 95% CrI estimated using the Cori et al. method and the estimate of reproduction number with 95% CI utilizing the GGM for simulating the parameters employed in the renewal equation method for the COVID-19 epidemic in Colombia as of March 27, 2020 (first 30 days). The red solid line is the mean reproduction number estimated from the renewal equation method and the red dashed lines are the 95% CI coverage around the mean reproduction number. The green solid line is the median instantaneous reproduction number estimated from the Cori method and the green dashed lines are the 95% CrI coverage around the median instantaneous reproduction.

**Fig 8.**
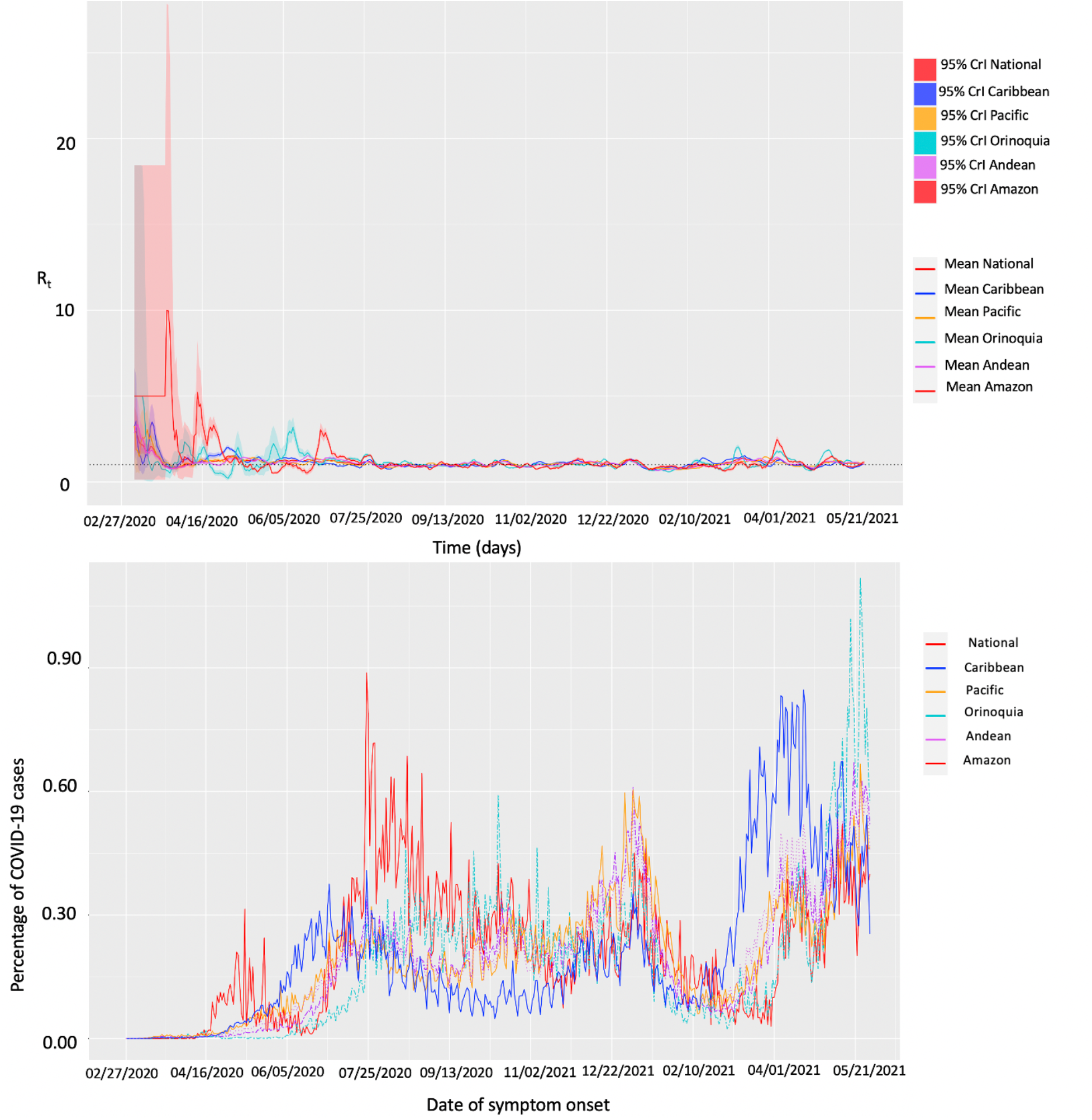
Upper panel: Instantaneous reproduction number with 95% credible intervals for the COVID-19 epidemic in Colombia as of May 30, 2021. The red solid line represents the mean reproduction number for Colombia and the red shaded area represents the 95% credible interval around it. The blue solid line represents the mean reproduction number for Caribbean region and the blue shaded area represents the 95% credible interval around it. The yellow solid line represents the mean reproduction number for Pacific region and the yellow shaded area represents the 95% credible interval around it. The cyan solid line represents the mean reproduction number for Orinoquía region and the cyan shaded area represents the 95% credible interval around it. The purple solid line represents the mean reproduction number for Andean region and the purple shaded area represents the 95% credible interval around it. The pink solid line represents the mean reproduction number for Amazon and the pink shaded area represents the 95% credible interval around it. Lower panel: The percentage of the COVID-19 cases in Colombia, nationally and regionally as of May 30, 2021. The red solid line represents the percentage of cases in Colombia, the blue solid line represents the percentage of cases in the Caribbean region, the yellow solid line represents the percentage of cases in the Pacific region, the cyan dotted line represents the percentage of cases in the Orinoquía region, the purple dashed line represents the percentage of cases in the Andean region and the pink dotted line represents the percentage of cases in the Amazon region.

At the regional level, the majority of the fluctuations in *R_t_* were observed during the first 130 days (until July 5, 2020) of the pandemic after which estimates of *R_t_* remain ∼1.0. For the Caribbean region the early estimate of reproduction number was *R_t_* ∼3.42 (95% CrI: 1.85-5.69) on March 6, 2020. The reproduction number peaked on March 16, 2020, with an estimate of *R_t_* ∼3.46 (95% CrI: 2.69-4.36) and declined thereafter to remain consistently above 1.00 (*R_t_* ∼1.00-3.21) until July 7, 2020. The reproduction number then fluctuated ∼1.00 until February 11, 2021. Since then, the estimates of *R_t_* have remained between ∼0.8-1.5 with the most recent estimate of *R_t_* ∼1.03 (95%CrI: 1.02-1.05) as of May 30, 2020. For the Pacific region, the early estimate of reproduction number as estimated on March 5, 2020 was *R_t_* ∼3.04 (95% CrI: 1.28-5.96). The reproduction number remained above 1.50 in the next 15 days. This was followed by the estimates of *R_t_* consistently above 1.00 until the end of July 20, 2020, after which *R_t_* fluctuated ∼1.00 until February 2021. From March to April 13, 2021, the estimates of *R_t_* remained between 0.9-1.5 with the most recent estimate of *R_t_* ∼1.05 (95% CrI: 1.04-1.06). The reproduction number in the Orinoquía region was estimated at *R_t_* ∼3.46 (95% CrI: 0.25-14.97) from March 5, 2020, to March 11, 2020, and declined thereafter to fluctuate ∼1.00 until the end of May 2020. From June 1, 2020, till the end of August 2020 the estimates of *R_t_* remained above 1.00 followed by fluctuations in *R_t_* around 1.00 with the most recent estimate of *R_t_* ∼1.01 (95% CrI: 0.98-1.03) as of May 30, 2021. The reproduction number in the Andean region peaked at *R_t_* ∼3.28 (95% CrI: 2.27-4.55) on March 6, 2020. This was followed by the estimates of reproduction number above 1.00 until July 27, 2020. Since then, *R_t_* has fluctuated ∼1.00 with the most recent estimate of *R_t_* ∼1.03 (95% CrI: 1.02-1.03) as of May 30, 2021. In contrast, the reproduction number in the Amazon region was estimated at *R_t_* ∼3.46 (95% CrI: 0.25, 14.9) on March 5, 2020. This was followed by the estimates of *R_t_* consistently above 1.00 until May 11, 2020 which have since fluctuated ∼1.00 with the most recent estimate of *R_t_* ∼1.16 (95% CrI: 1.11-1.21) as of May 30, 2021 (Fig 8). As can be observed for national as well as the regional level data, the Cori method tends to overestimate *R_t_* early in the time series as infections occurring before the first date in the time series remain missing terms in the denominator [83].

### Estimate of reproduction number, R from genomic data analysis

Between February and April 2020, 136 analyzed Colombian sequences were sampled. These sequences were spread along the whole global SARS-CoV-2 phylogeny (Fig 9) and split into multiple clusters. This indicates multiple introductions of SARS-CoV-2 to the country during the initial pandemic stage (February 27-April 5, 2020). For the largest cluster of size 35, growth rate for the early ascending phase of the COVID-19 pandemic in Colombia was estimated to be ∼12.718. Subsequently, the reproduction number was estimated at, R∼1.2 (95% HDP interval: [1.03, 1.44]), indicating sustained disease transmission during the early phase of the pandemic.

### Spatial analysis

Fig 10 shows the result from pre-processing COVID-19 data into growth rate functions. The results of clustering are shown in Fig 11 as a dendrogram plot. The four predominant clusters identified include the following departments and districts:

**Fig 9.**
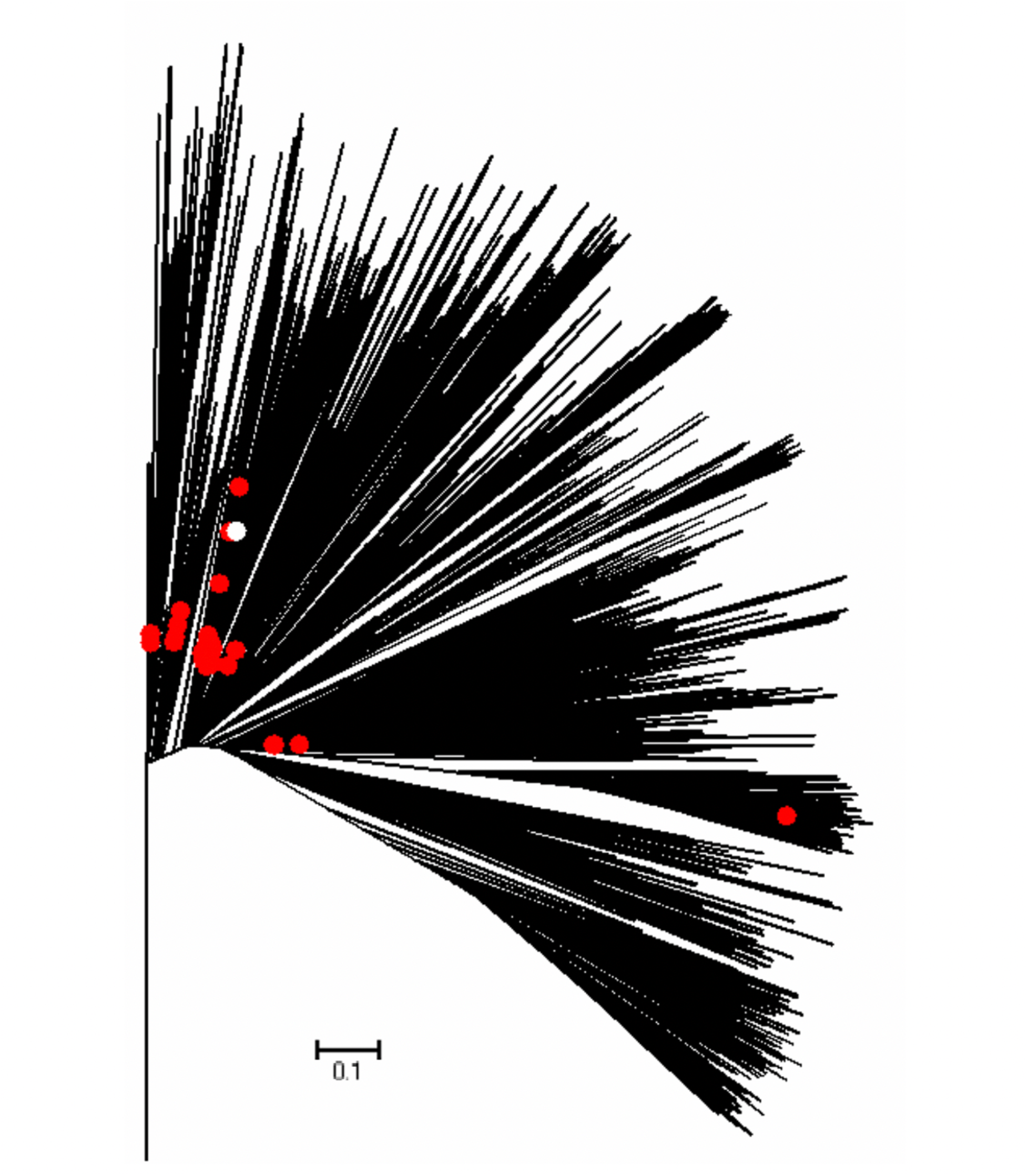
Global ML tree for SARS-CoV-2 genomic data for Colombia.

**Fig 10.**
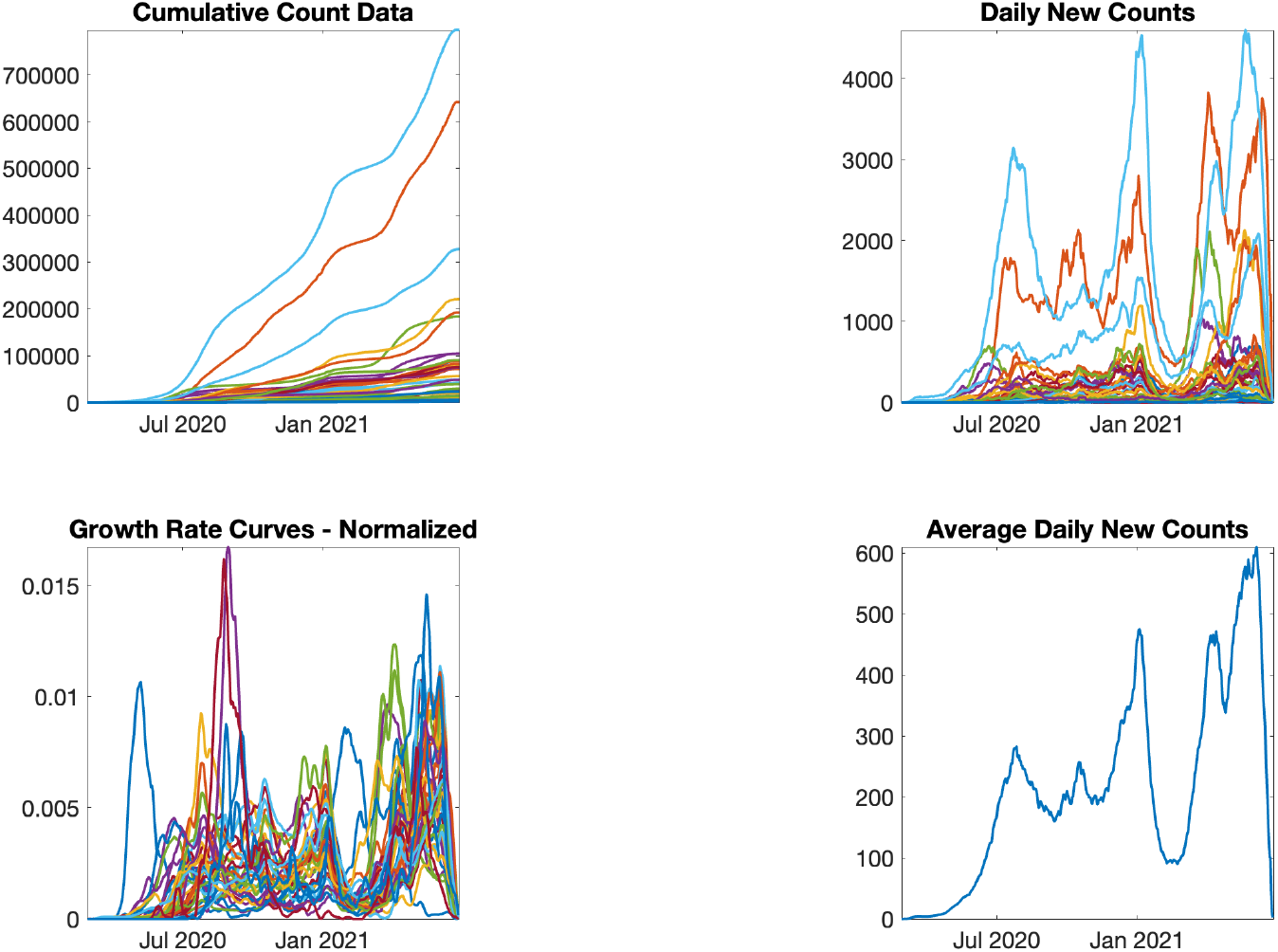
Pre-processing COVID-19 data into incidence rate functions. From left to right: original lab-confirmed COVID-19 cases, curve of daily new cases, smoothed and scaled rate curves, average of rate curves before scaling and smothing.

**Fig 11.**
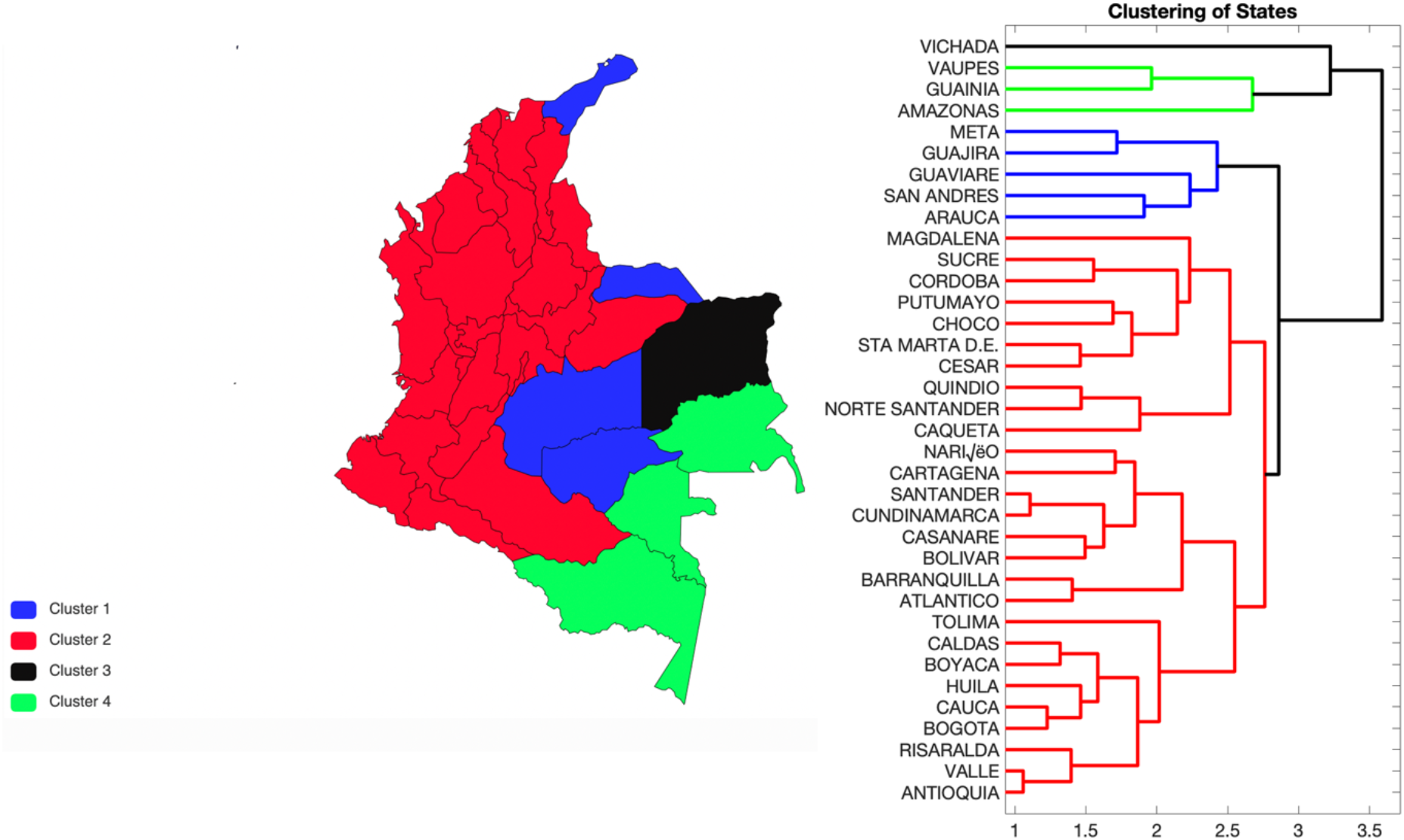
Clustering of states according to the shapes of their rate curves. Cluster 1 is shown in blue, cluster 2 is shown in red, the smallest cluster, cluster 3 is shown in black whereas cluster 4 is shown in the green. One can see that states with similar shapes of rates curves are geographically close to each other. The left panel shows the geographic distribution of the clusters and the right panel shows the dendrogram.

Cluster 1: Arauca, Guajira, Guaviare, Meta, San Andres

Cluster 2: Antioquia,, Atlántico, Barranquilla D.E., ., Bolivar, Boyacá, Bogota D.C., Caldas, Caquetá, Cartagena D.T y C., Casanare, Cauca, Cesar, Choco, Cordoba, Cundinamarca, Huila, Magdalena, Nariño, Norte Santander, Putumayo, Quindío, Risaralda, Santander, Santa Marta D.T y C, Sucre, Tolima, Valle del Cauca

Cluster 3: Vichada

Cluster 4: Amazonas, Guainía, Vaupes

Fig 12 shows the average shape of incidence rate curves in each cluster and the overall average. Fig 13 shows the mean growth rate curves and one standard-deviation bands around it, in each cluster. The average incidence patterns in these four clusters are very distinct and clearly visible (Fig 13). For cluster 1, the rate increases rapidly from April to July 2020 and then declines to increase again at the end of November 2020 followed by another incline at the start of January 2021. For cluster 2, we can observe a three modal growth pattern. There is rapid increase in growth rate in July 2020 followed by a decline and continuous fluctuations. This pattern is followed by two more peaks, one in January 2021 and the second bimodal peak can be observed in March and April 2021. Cluster 3 shows a slow growth rate until July 2020 followed by a rapid rise in growth curve until mid-September 2020. This is followed by a gradual decline of the growth curve with fluctuations and another large peak in June 2021. For cluster 4, the slow growth rate is followed by a spike in September 2020 and multiple fluctuations. This is followed by another peak in June 2021 (Fig 13).

**Fig 12.**
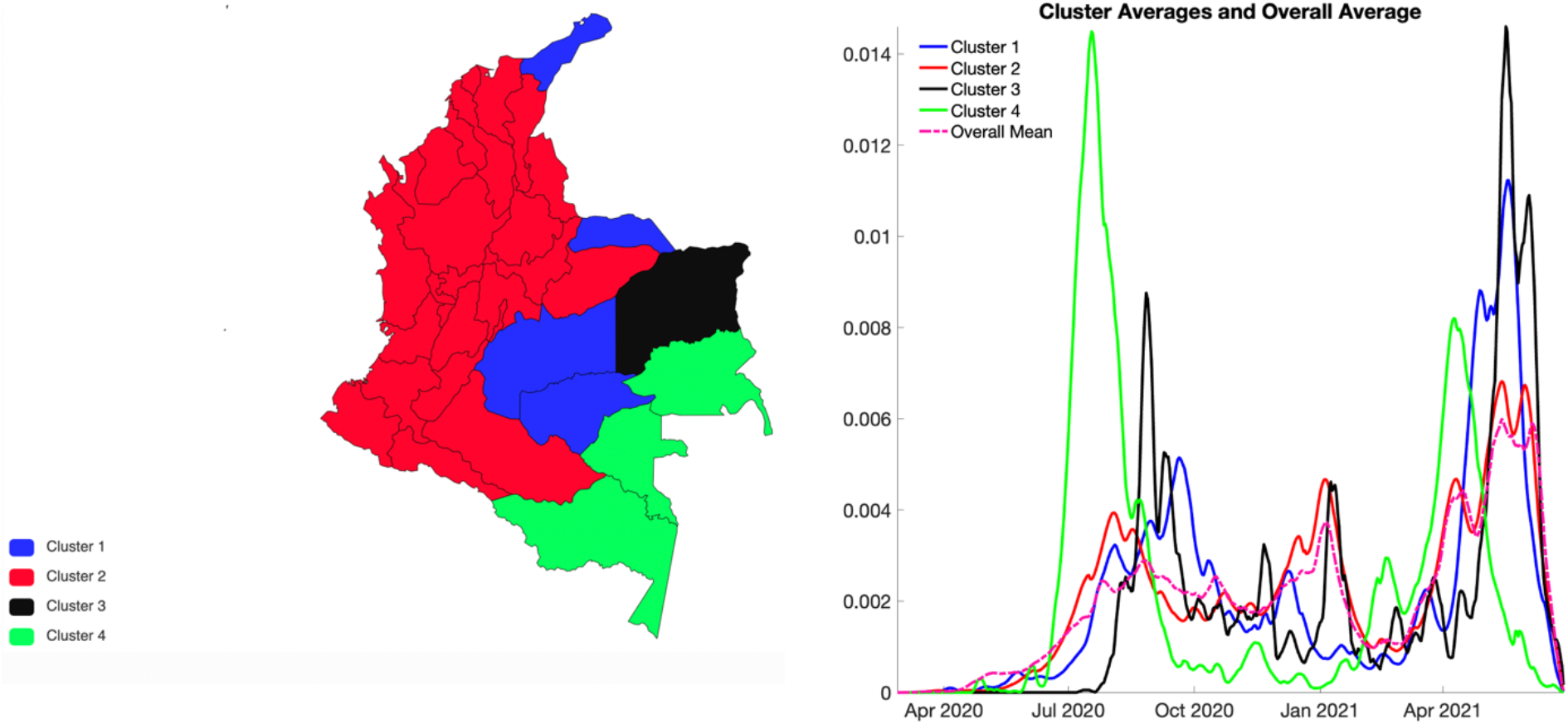
Clusters of states by their growth rates. Cluster 1 is shown in blue, cluster 2 is shown in red, cluster 3 is shown in black, cluster 4 is shown in green and the overall cluster mean in is shown in pink curve. The left panel shows the geographic distribution of the clusters and the right panel shows the average growth rate curves for each cluster (solid curves) and their overall average (pink broken curve).

**Fig 13.**
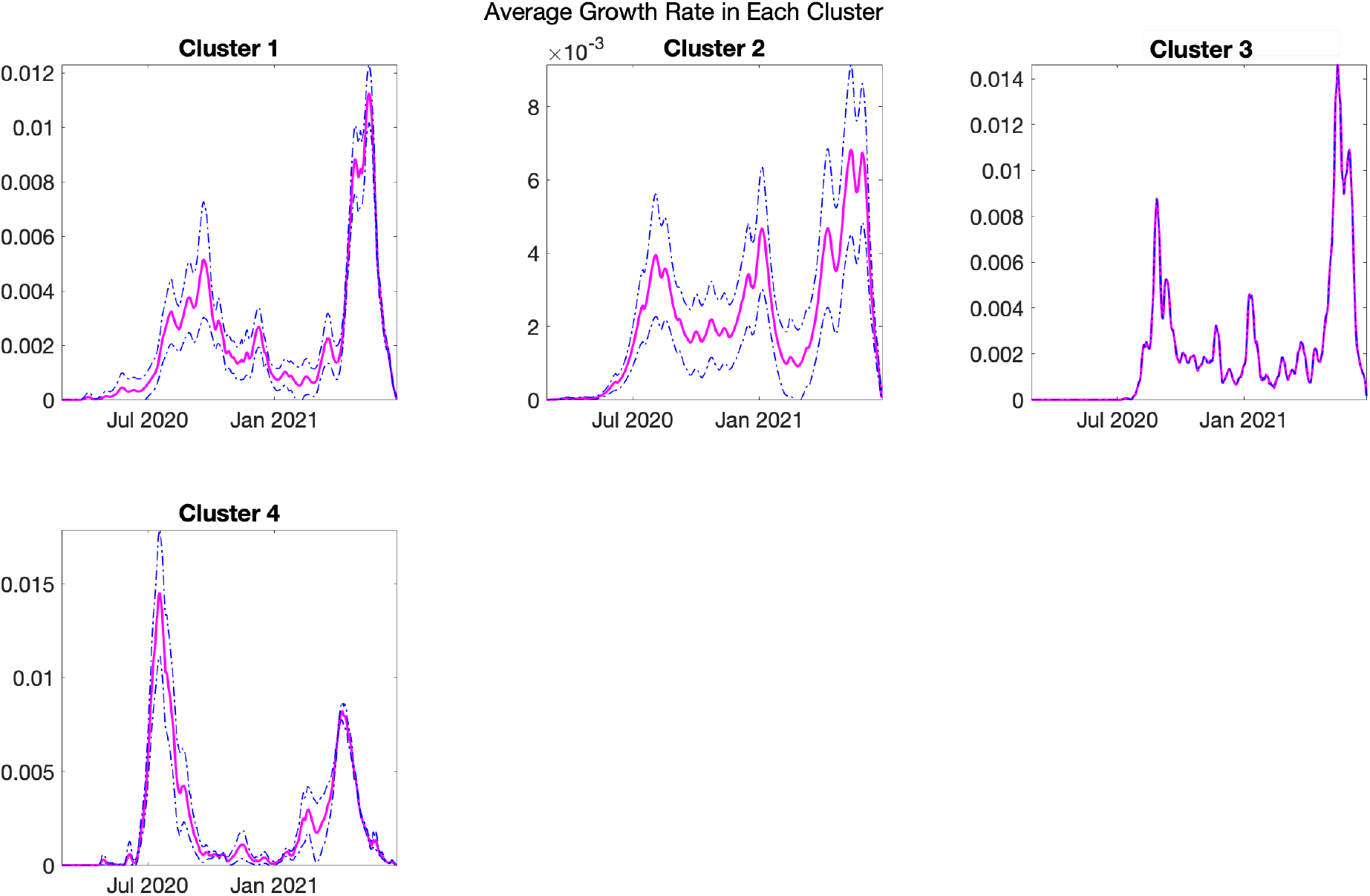
Average shapes of the COVID-19 incidence rate curves, along with a one standard-deviation band around the average, in each of the clusters.

### Mobility data

The curve of mobility trends from Apple tracked in the form of driving and walking declined in April 2020 to less than 60% corresponding to multiple interventions focused on the social distancing mandates as implemented by the government of Colombia. The mobility then started to increase in June and peaked in July 2020. Soon after a decline in mobility was observed in August and September 2020 which was followed by fluctuations in mobility trends around the baseline until the end of June 2021 (Fig 14). Correspondingly it can be observed that as the mobility increased in July 2020, the number of cases also peaked after which the cases fluctuated at a steady level (∼7000 cases per day) until the end of December 2020.

**Fig 14.**
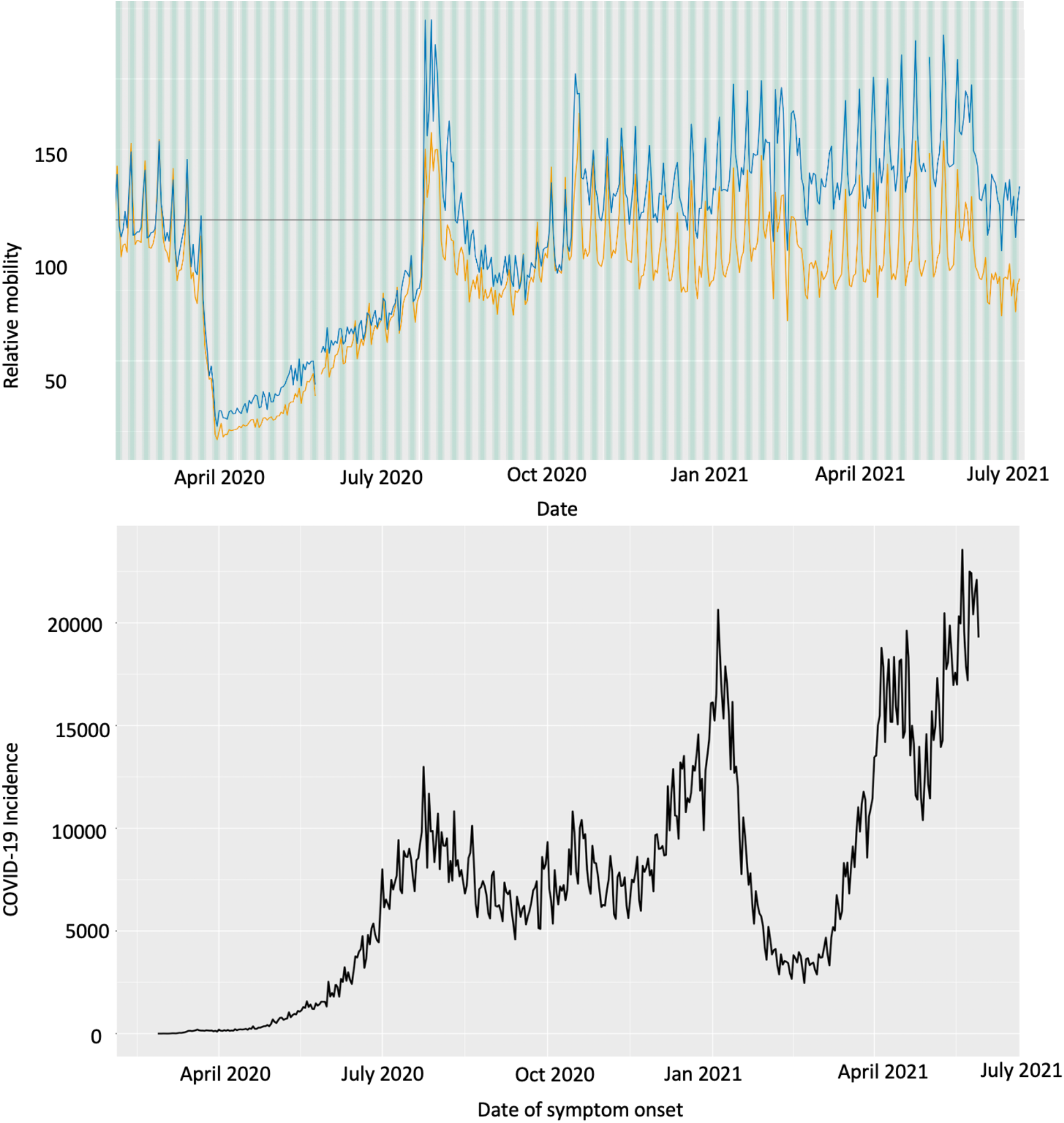
Upper panel: Apple mobility trends data showing the variation in walking and driving among the population in Colombia. Orange curve shows the driving trend and the blue curve shows the walking trend. Lower panel: COVID-19 incidence curve in Colombia by the dates of symptoms onset as of June 30, 2021.

The curves of mobility from Google data tracked in the form of visits to retail and recreation, grocery, pharmacy, parks, transit stations and workplaces all follow the same pattern; declining in April 2020 to less than 60% incoherence with the Apple mobility data and corresponding to the government’s interventions to contain the spread of the virus. The mobility started to increase gradually thereafter but remained below the baseline until December 2020. The greatest fluctuations in mobility were observed in January 2021, the same time when the cases also peaked. The mobility has since fluctuated around the baseline. The residential category is measured in duration rather than the number of visitors, so we do not compare it with other 5 categories. Moreover, since people spend a larger amount of time at home, changes in the residential category remain inconspicuous (Fig 15).

**Fig 15.**
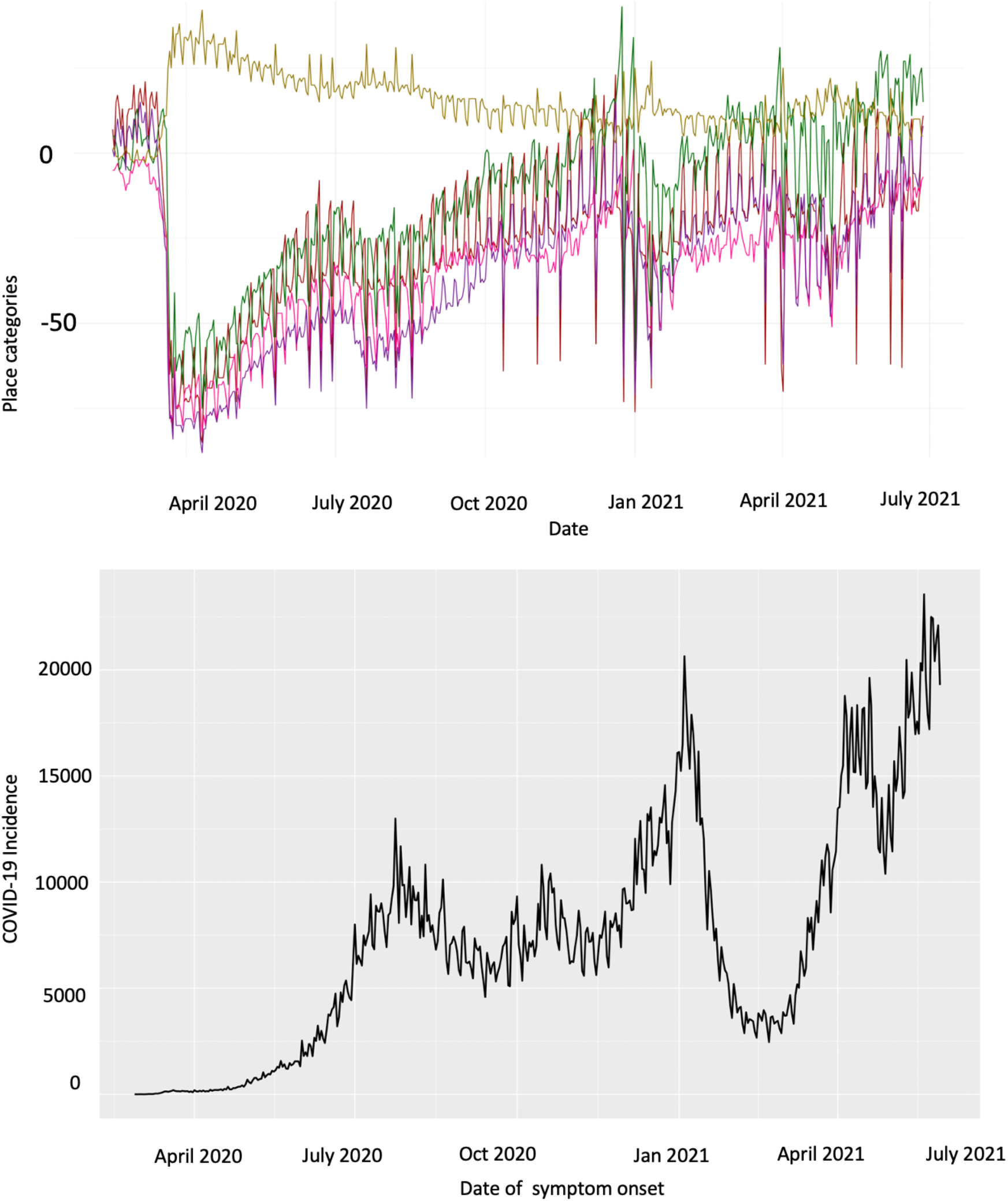
Upper panel: Google mobility trends data showing the variation in movement of people across the following six categories: retail and creation (maroon solid curve), grocery and pharmacy (green solid curve), parks (pink solid curve) and transit stations (purple solid curve), workplaces (red solid curve) and residential (golden solid curve) among the population in Colombia. Lower panel: COVID-19 incidence curve in Colombia by the dates of symptoms onset as of June 30, 2021. Lower panel: COVID-19 incidence curve in Colombia by the dates of symptoms onset as of June 30, 2021.

### Twitter data analysis: social media trends

The epidemic curve for Colombia is overlaid with the curve of tweets indicating the stay-at-home orders in Colombia. In Colombia the engagement of people with the #quedateencasa hashtag (stay-at-home order hashtag) fluctuated at a steady level during the course of the pandemic with a slight decline in the frequency of tweets observed after June 2020 while the case counts continued to increase, which could point towards the apathy or frustration of the public towards the lockdowns and restrictions. This could also imply that the population does not follow the government’s stay-at-home orders. The decline in the number of tweets indicating stay-at-home orders was followed by a sharp increase in the frequency of tweets indicating the stay-at-home order in November 2020 which gradually declined until April 2021 and has since fluctuated at a steady level, ∼1000 tweets a day. On the contrary, the cases peaked in July 2020 levelled off ∼7000 cases per day until December 2020 (Fig 16). Another peak in case incidence was observed in January 2021 followed by a sharp decline in case incidence. This was followed two upsurges in case incidence in November 2020 and April 2021. The correlation coefficient between the epidemic curve of cases by dates of onset and the curve of tweets representing the stay-at-home orders was estimated at R=0.1095 from March 12, 2020 to June 29, 2021.

**Fig 16.**
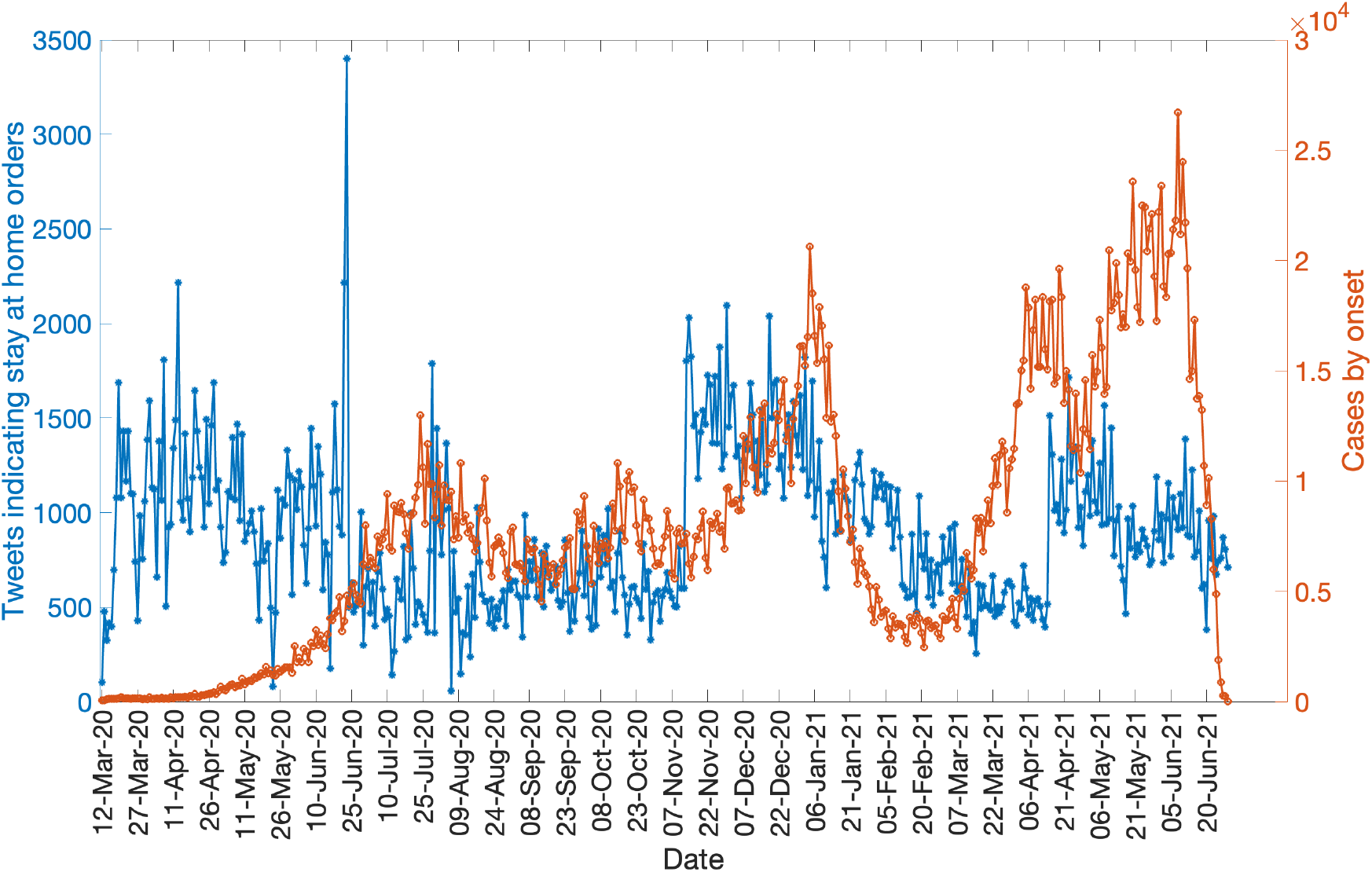
The daily number of tweets indicating stay at home orders (blue) and the daily number of new COVID-19 cases by dates of symptoms onset (orange) as of June 30, 2021

## Discussion

In this study, we explore the transmission dynamics of the COVID-19 pandemic in Colombia by applying multiple methodologies to analyze five data sources. Our model calibration and forecasting indicate that the sub-epidemic model outperforms the GLM and Richards growth model in terms of three calibration performance metrics at the national and the regional levels in Colombia. However, the forecasting performance metrics do not yield one single model as the best model. More importantly, the regional and national level forecasts point towards a declining trend in the epidemic trajectory except for the GLM which predicts an upward trend for the Andean, Amazon, Pacific and Orinoquía regions and the sub-epidemic model which predicts an upsurge in case counts for the national and Andean region. Estimation of national and regional reproduction numbers show a sustained disease transmission during the early phase of the COVID-19 pandemic exhibiting sub-exponential growth dynamics. Moreover, the estimates of reproduction number from the case incidence data during the most recent months show fluctuation of *R_t_* ∼1.00 without any major surges. Estimates of *R*_0_ from the genomic data agree with those obtained from the case incidence data. Spatially we can divide the Colombian departments into four distinct clusters based on the growth rate patterns in each cluster. Moreover, while the mobility in Colombia increased in June and July 2020, corresponding to the observed peak in case incidence in July 2020, a spike in the number of tweets indicating the stay-at-home orders was observed in November 2020 when the case incidence had levelled off.

The scaled incidence of daily COVID-19 new cases by the date of symptom onset for all regions and the national data shows multimodal peaks as of May 30, 2021 (Fig 8). As observed the national COVID-19 epidemic peaked in mid-July 2020 followed by a decline in case incidence in mid-September 2020, only to rise again. The national curve declined at the beginning of year 2021. However, April 2021 showed another surge in the COVID-19 cases with estimates of *R_t_* ∼1.00, indicating continued virus transmission in the country.

The early transmission dynamics of SARS-CoV-2 exhibit similarity at the national and regional levels. The COVID-19 pandemic in Colombia exhibited sub-exponential growth dynamics (0<*p*<1.0) during the ascending phase of the outbreak at the national and regional levels. The sub-exponential growth pattern of the COVID-19 pandemic in Colombia can be attributed to a myriad of factors including non-homogenous mixing, spatial structure, population mobility, behavior changes and interventions [84]. Our results are consistent with the sub-exponential growth patterns of COVID-19 outbreaks observed in other Latin American countries including Mexico [85] and Chile [86]. Moreover, the estimates of early transmission potential (*R_t_*) indicate sustained disease transmission in Colombia at the national and regional levels. In this context, we observed that these results are compatible with the estimates of early reproduction numbers retrieved from other countries, including Peru, Chile, Brazil and Mexico which followed similar COVID-19 outbreaks around the same time period [87–90].

The overall estimates of transmission potential (instantaneous reproduction number) throughout the COVID-19 pandemic in Colombia displayed substantial spatiotemporal variation. Indeed multiple factors can be attributed to influence the reproduction number including the transmissibility of an infectious agent, individual contact rates, individual susceptibility and mitigation strategies [57, 91]. Our results indicate that the national reproduction number remained substantially high (*R_t_*>1) until the end of July 2020, after which it declined and fluctuated around ∼1.0 with no major deviations. The most recent estimates of *R_t_* indicate second surge in reproduction number with estimates of *R_t_* greater 1.0 as of May 30, 2021. At the regional level, Orinoquía, Pacific and Caribbean regions displayed the greatest fluctuation in the transmission potential. Moreover, the estimates of *R_t_* remained between 3.02-3.46 until March 6, 2020, for all regions. As the pandemic progressed and control interventions were put in place, the estimates of *R_t_* declined to fluctuate ∼1.0 around. The estimates of *R_t_* remained consistently above 1.00 in the Caribbean, Andean and Pacific regions until July 2020, after which *R_t_* fluctuated ∼1.00. The results of our genomic analysis for Colombia also indicate sustained disease transmission in the country during the early ascending phase of the pandemic with *R*_0_>1.00.

In general, Colombia has observed sustained disease transmission of SARS-CoV-2 despite the implementation of social distancing interventions. As can also be seen from Twitter analysis, the number of cases by dates of onset are positively correlated with the tweets indicating the stay-at-home orders. A plausible explanation of this phenomenon could be that the people might have stopped following the government’s preventive orders to stay at home as a result of pandemic fatigue or due to confusion regarding the national and regional policies. There was a decline in the mobility profile in Colombia after the implementation of social distancing mandates and declaration of national emergency. However, as the pandemic progressed, the mobility of the public reached a normal level with fluctuations around the baseline. A study reported that the average mobility reduction in Colombia fluctuated during the pandemic, starting at ∼80% in April 2020, and reaching ∼30% by the end of September 2020 [92]. In comparison Argentina and Chile have reported the most steady mobility reductions of ∼80% and 70% respectively during the course of the pandemic [92].

At level of each administrative division, we analyze the spatial-temporal dynamics of the COVID-19 pandemic in Colombia. Our classification of epidemic patterns in Colombia at the department level, including their 4 main districts, shows distinct variations in growth rates across different departments. For instance, cluster 1 including Arauca, Guajira, Guaviare, Meta, San Andres shows a rising pattern in growth rate. In contrast, cluster 3 that includes Vichada and cluster 4 that includes Amazonas, Guainía, Vaupes show stable growth at lower rates. This information can be utilized by the departments, districts, and regions in guiding their decision regarding the implementation of public health measures. For example, administrative divisions in cluster 1 show an early increasing growth pattern and may need strict public health measures to contain the epidemic.

Appropriate short-term forecasts at the national and regional levels can also help guide the intensity and magnitude of public health interventions required to contain the epidemic. In general, the national and regional level short-term forecasts from the Richards model point towards a sustained decline in the over-all case counts. However, the GLM forecasts show an increasing trajectory of the COVID-19 pandemic for the Andean, Pacific and Orinoquía region. Similarly, the sub-epidemic wave model’s forecast for the national and Andean region shows an upward trend in the trajectory of the COVID-19 pandemic and a steady trend in the trajectory of the COVID-19 pandemic for the Amazon and Caribbean regions. As with the sub-epidemic wave model, it captures the underlying structure of the sub-epidemic building blocks of the coarse scale epidemic. The three models generally provide a good fit to the COVID-19 case counts for the most recent part of the epidemic. However, in our analysis the sub-epidemic model performs better than the GLM and Richards model in the calibration periods. However, for the forecasting periods, no model can be declared as the best model. The Richards model performs better for the Amazon and Andean region, GLM performs better for the Caribbean region and the sub-epidemic model performs better for the national, Pacific and Orinoquía regions.

This study has some limitations. First, we excluded the last 30-day case counts from our study in order compare the performance of our models in the forecasting phase. Moreover, excluding the last 30 data points allows us to overcome the influence of reporting delays on our analysis. Specifically, we utilize the case counts based on the dates of onset in this study, because delays in case reporting, testing rates and factors related to the surveillance systems can influence our epidemic projections. Secondly, we relied on the daily updates of cases in the official surveillance system of Colombia, which can sometimes underreport the cases. Third, the phenomenological models applied in this study do not explicitly account for behavioral changes, and thus the results such as the predicted decline or stability in the epidemic trajectory should be interpreted with caution. Fourth, some authors [83] shifted the dates of symptom onset backwards to approximate the date of infection when applying the EpiEstim package to estimate the instantaneous reproduction number. However, in this study we stick to the date of symptoms onset for the sake of comparison across methods to estimate the reproduction number. Lastly, the unpredictable social component of the epidemic on ground was also a limiting factor for the study as we did not know the ground truth epidemic pattern when the forecasts were generated.

In conclusion, the country observed a surge in case counts as the mobility increased in July 2020; however, since the end of 2020, the national and regional reproduction numbers have fluctuated around ∼1.00 indicating sustained virus transmission in the region with estimates of *R_t_* > 1.00 as of May 30, 2021. Moreover, the population of Colombia has had mixed reactions towards the stay-at-home orders as can be inferred from the Twitter analysis, contributing to the virus transmission in the country. Importantly, the spatial analysis indicates that departments like Arauca, Guajira, Guaviare, Meta, San Andres need stronger public health strategies to contain the rising patterns in growth rates. Although Richards model point towards a sustained decline in the case counts in general, the sub-epidemic model indicates an upward trend in case incidence for the national and Andean regions and the GLM shows an upsurge in case counts for the Pacific, Orinoquía and the Amazon regions. Hence, the forecasts need to be interpreted with caution given the spatial heterogeneity in transmission rates as well as dynamic implementation and lifting of the social distancing measures. The phenomenological growth models employed in this study to forecast and estimate reproduction numbers are particularly valuable for providing rapid predictions of the epidemics in complex scenarios that can be used for real-time preparedness because these models do not require specific disease transmission processes to account for the interventions.

## Author contributions

Conceptualization, G.C. and A.T.; methodology, G.C, A.T., P.S., J.M.B., A.S.; validation, G.C.; formal analysis, A.T., G.C.; investigation, A.T., G.C, J.M.B., A.E.D.; resources, G.C., A.T. J.M.B., P.S. A.S.; data curation, A.T.; writing—original draft preparation, A.T.; writing, review and editing, A.T., G.C., A.E.,A.S., O.P., S.D., S.O., T.C., X.H., J.M.B., A.E.D., P.S., R.L, L.Y.L.D, R.B., I.C-H.F., E.S., A.K., A.S.; visualization, A.T., G.C; supervision, G.C.; project administration, G.C.; funding acquisition, G.C., P.S., R.B, A.T., L.Y.L.D., E.S. All authors have read and agreed to the published version of the manuscript.

## Data Availability

Case incidence data by the dates of symptom onset is available from the Colombian
Ministry of Health. url: https://coronaviruscolombia.gov.co/Covid19/index.html.
Genomic data is available from the global initiative on sharing avian influenza data
(GISAID) repository. url: https://www.gisaid.org/
Apple mobility data is available from the Apple's mobility trends reports. url:
https://covid19.apple.com/mobility.
Google mobility data is available from the Google mobility page (COVID-19 Community
Mobility Reports Google 2020)
Twitter data is available from the Twitter data set of the COVID-19 chatter.

https://www.gisaid.org/

https://covid19.apple.com/mobility

