## Supplemental file 1 for "An investigation of spatial-temporal patterns and predictions of the COVID-19 pandemic in Colombia, 2020-2021"

**Online Supplementary Materials**

**Amna Tariq*^1^, Tsira Chakhaia^1^ , Sushma Dahal^1^, Alexander Ewing^1^, Xinyi Hua^2^ , Sylvia K. Ofori^2^ , Olaseni Prince^1^, Argita Salindri^1^ , Ayotomiwa Ezekiel Adeniyi^3^ , Juan M. Banda^3^, Pavel Skums^3^, Ruiyan Luo^1^, Leidy Y. Lara D**$\acute{\mathbf{i}}$**az^4^, Raimund Bürger^4^, Isaac Chun-Hai Fung^2^, Eunha Shim^5^, Alexander Kirpich^1^, Anuj Srivastava^6^, Gerardo Chowell^1^**

**Model descriptions**

(i) **Generalized logistic growth model**

The generalized logistic growth model (GLM) [1] displays a range of epidemic growth patterns including the polynomial and exponential growth patterns. GLM characterizes epidemic growth by estimating three parameters: (i) the intrinsic growth rate, *r* (ii) a dimensionless "deceleration of growth" parameter, *p* ∈[0,1] and (iii) $k_{0},$representing the final epidemic size [1]. The varied epidemic growth patterns are observed by the modulation of deceleration of growth parameter resulting in the exponential growth dynamics (*p*=1), sub-exponential growth (0<*p*<1), or constant incidence (*p*=0) patterns, if $k_{0}=\infty$. When $k_{0}<\infty$ and *p*=1, the GLM is the simple logistic growth model. The following differential equation gives the GLM model:

$$\frac{dC(t)}{dt}=rC(t)^{p}\left( 1-\frac{C\left( t \right)}{k_{0}} \right) , (1)$$

where 𝐶(𝑡) denotes the cumulative number of cases at time 𝑡, and ${dC(t)}/{dt}$ describes the incidence at time *t* [1].

(ii) **Richards growth model**

The well-known Richards model [2] is a simple extension of the logistic model that relies on three parameters; growth rate, 𝑟, final epidemic size, $k_{o}$ and the scaling parameter, *a.* The scaling parameter, *a,* measures the deviation from the symmetric S-shaped dynamics exhibited by the simple logistic growth curve [2-4]. The following differential equation gives the Richards model:

$\frac{dC\left( t \right)}{dt}=rC(t)\left[ 1-\left( \frac{C(t)}{k_{o}} \right)^{a} \right]$,

where 𝐶(𝑡) represents the cumulative case count at time 𝑡. We remark that the Richards growth model has the explicit solution

$C\left( t \right)=\frac{k_{0}C\left( 0 \right)\exp(rt)}{k_{0}^{a}+C(0)^{a}(\exp\left( art \right)-1{))}^{1/a}}$,

while the GLM does not admit a closed-form solution. For a unified treatment of all phenomenological growth models, we always refer to the corresponding differential equation in each case irrespective of the existence of a closed-form solution. Details are provided in a prior study [5].

(iii) **Sub-epidemic wave model.**

The sub-epidemic model [6] is based on the premise that various profiles of overlapping sub-epidemics shape the aggregated reported epidemic wave. In particular, this modeling approach supports complex temporal dynamic patterns, such as oscillating dynamics leading to damped oscillations or endemic states. This model characterizes each group sub-epidemic utilizing a three-parameter generalized logistic growth model as explained above and given in equation (1).

We model an epidemic wave comprising of *n* overlapping sub-epidemics using a system of coupled differential equations, as follows:

$$\frac{dC_{i}(t)}{dt}= rA_{i-1}\left( t \right)C_{i}(t)^{p}\left( 1-\frac{C_{i}(t)}{k_{i}} \right)$$

In this equation, $C_{i}(t)$ describes the cumulative cases for the i^th^ sub-epidemic, and $k_{i}$ is the size of sub-epidemic *i* (*i*=1,2,….*n*). Parameters *r* and *p* are the same across the sub-epidemics. The coefficient $A_{i}(t)$ is an indicator variable that models the onset timing of (*i*+1)^th^ sub-epidemic, making sure that sub-epidemics comprising an epidemic wave follow a regular structure. Therefore,

$A_{i}\left( t \right)=\left\{ \begin{aligned} 1 C_{i}\left( t \right)>C_{thr} \\ 0 Otherwise \end{aligned} \right. i=1,2,3,\ldots n$,

Where $1\leq C_{thr}<k_{o}$ and $A_{1}\left( t \right)=1$ for the first sub-epidemic. Therefore, when *n* = 1 and *p* = 1, the sub-epidemic model becomes the simple logistic model. Moreover, for the subsequently occurring sub-epidemics, the size of *i*^th^ sub-epidemic ($k_{i})$ declines exponentially at a rate q:

$k_{i}=k_{0}e^{-q(i-1)}$,

where $k_{0}$is the final size of the epidemic (when the epidemic ends). The exponential decline in the size of *i*^th^ sub-epidemic can occur due to multiple factors, including the effect of interventions, changes in disease transmission dependent on seasonality and behavior changes.

**(iv) Generalized growth model (GGM)**

The generalized growth model (GGM) characterizes the early ascending phase of the epidemic by estimating two parameters: (1) the intrinsic growth rate, $r$; and (2) a dimensionless “deceleration of growth” parameter, 𝑝. The deceleration of growth parameter, 𝑝, allows this model to capture a range of epidemic growth profiles. The following differential equation gives the GGM model:

$$\frac{dC(t)}{dt}=C^{'}\left( t \right)=rC(t)^{p}$$

In this equation, $C^{'}(t)$ describes the incidence curve over time $t$, solution 𝐶(𝑡) describes the cumulative number of cases at time 𝑡 and 𝑝 is the modulating "deceleration of growth" parameter ($0\leq p\leq$1). This equation displays constant incidence over time if 𝑝=0 and becomes an exponential growth model for cumulative cases if 𝑝 =1. The model shows sub-exponential growth dynamics if 𝑝 is in the range 0< 𝑝 <1 [3, 7].

**Model calibration and forecasting approach**

We estimate the best-fit solution for each model using non-linear least squares fitting procedure for each of the three models (i.e., the GLM, Richards growth model and the sub-epidemic wave model) [8]. This process yields the best set of parameter estimates $\hat{\Theta}$ *= (θ_1_, θ_2_, …, θ_m­_),* where *m* is the number of parameters of interest, by minimizing the sum of squared errors between the model fit, $f(t,\hat{\Theta})$and the data estimates, $y_{t_{i}}$. The parameters $\hat{\Theta}=argmin\sum_{t=1}^{n} (f(t,\hat{\Theta}{-y_{t_{i}})}^{2}$define the best fit model $f\left( t,\Theta\right)$. In this analysis, $\hat{\Theta}=(r,p,k_{o},q and C_{thr})$ corresponds to the set of parameters of the sub-epidemic model, $\hat{\Theta}=(r,k_{0},a)$ corresponds to set of parameters of the Richards model and $\hat{\Theta}=(r,p,k_{0})$ corresponds to the set of parameters of the GLM model [3]. For the sub-epidemic wave model, we determine the initial best guesses of parameter estimates. However, for the GLM and Richards growth model we initialize the parameter estimates for the nonlinear least squares method [8] over a wide range of plausible parameters from a uniform distribution using Latin hypercube sampling [9]. This allows us to test the uniqueness of the best model fit. The initial conditions are set at the first data point for each of the three models [3].

Uncertainty bounds around the best-fit solution are generated using a parametric bootstrap approach with replacement of data, where we assume a negative binomial error structure for the sub-epidemic model. A negative binomial error structure is also used to generate the uncertainty bounds of the Richards growth model and the GLM. For both these models, variance is assumed to be 250.49 times of the mean for national data, 8.50 times of the mean for Amazon region, 143.27 times of the mean for Andean region, 162.21 times of the mean for the Caribbean region, 28.54 times the mean for the pacific region, and 30.69 times of the mean for the Orinoquia region. This variance is based on the noise in the data and calculated by averaging mean to variance ratio obtained from the data. A detailed description of this method is provided in a prior study [3].

Each of the *M*=300 best-fit parameter sets is used to construct the 95% confidence intervals for each parameter by refitting the models to each of the *M* datasets generated by the bootstrap approach during the calibration phase. Further, each of the *M* best-fit model solutions is used to generate *m*= 30 additional simulations with a Poisson error structure for the sub-epidemic model and negative binomial error structure for the GLM and Richards model extended through a 30-day forecasting period. Finally, we construct the 95% prediction intervals using the 9000 (*M × m*) curves for the forecasting period. A detailed description of the parameter estimation methods can be found in previous literature [3, 10, 11].

**Performance metrics**

We utilize the following four performance metrics to assess the quality of our model fit and the 30-day ahead short-term forecasts: the mean absolute error (MAE) [12], the mean squared error (MSE) [13], the coverage of the 95% prediction intervals (95% PI) [13], and the mean interval score (MIS) [13] for each of the three models: GLM, Richards model and the sub-epidemic model.

We compare the model fit to the incidence data fitted to the model for evaluating the calibration performance. In contrast, we compare our forecasts with the incidence data for the forecast period for evaluating forecast performance.

The MSE and MAE assess the average deviations of the model fit to the observed data. The MAE is given by

$MAE=\frac{1}{n}\sum_{i=1}^{n} |f\left( t_{i},\hat{\Theta} \right)-y_{t_{i}}|$.

The MSE is given by

$MSE=\frac{1}{n}\sum_{i=1}^{n} (f\left( t_{i},\hat{\Theta} \right)-y_{t_{i}})^{2}$.

In both these equations, $y_{t_{i}}$is the time series of cases by date of onset, $t_{i}$ is the time stamp and $\hat{\Theta}$ is the set of model parameters. For the calibration period, *n* equals the number of data points used for calibration, and for the forecasting period, *n =* 30 for the 30-day ahead short-term forecast.

Moreover, in order to assess the model uncertainty and performance of PI, we used the 95% PI and MIS. The prediction coverage is defined as the proportion of observations that fall within the 95% PI and is calculated as

$PI coverage=\frac{1}{n}\sum_{i=1}^{n} \boldsymbol{I}\left\{ y_{t_{i}}>L_{t_{i}} \cap y_{t_{i}}<U_{t_{i}} \right\}$,

where $y_{t_{i}}$ is the case incidence data, $L_{t_{i}}$and $U_{t_{i}}$are the lower and upper bounds of the 95% PIs, respectively, *n* is the length of the period, and I is an indicator variable that equals 1 if value of $y_{t_{i}}$ is in the specified interval and 0 otherwise.

The MIS addresses the width of the PI as well as the coverage. The MIS is expressed as

$MIS= \frac{1}{n}\sum_{i=1}^{n} \left( U_{t_{i}}-L_{t_{i}} \right)+\frac{2}{0.05}(L_{t_{i}}-y_{t_{i}})I\left\{ y_{t_{i}}< \right.\left. L_{t_{i}} \right\}+\frac{2}{0.05} (y_{t_{i}}-U_{t_{i}})I\left\{ y_{t_{i}} \right.>\left. U_{t_{i}} \right\}$.

In this equation $L_{t_{i}}$, $U_{t_{i}}$, $y_{t_{i}}$*, n* and I are specified above for PI coverage. Therefore, if the PI coverage is 1, the MIS is the average width of the interval across each time point. For two models with equivalent PI coverage, a lower MIS value indicates narrower intervals [13].
